# Variant curation of the largest compendium of *FOXL2* coding and non-coding sequence and structural variants in BPES

**DOI:** 10.64898/2026.02.24.25339471

**Authors:** Charlotte Matton, Julie Van De Velde, Marieke De Bruyne, Stijn Van De Sompele, Sally Hooghe, Hannes Syryn, Miriam Bauwens, Eva D’haene, Annelies Dheedene, Martine Cools, Shoko Komatsuzaki, Ewelina Preizner-Rzucidło, Alison Ross, Christine Armstrong, Wendy Watkins, Andrew Shelling, Andrea L Vincent, Catherine Cassiman, Sascha Vermeer, David J Bunyan, Hannah Verdin, Elfride De Baere

**Affiliations:** Department of Biomolecular Medicine, Ghent University, Ghent, Belgium; Center for Medical Genetics Ghent, Ghent University Hospital, Ghent, Belgium; Department of Pediatric Endocrinology, Ghent University Hospital, Ghent, Belgium; Department of Internal Medicine and Pediatrics, Ghent University, Ghent, Belgium; Institute of Human Genetics, University of Würzburg, Biozentrum Am Hubland, Würzburg, Germany; Department of Molecular Genetics, Institute of Pediatrics, Jagiellonian University Medical College, Krakow, Poland; North of Scotland Regional Genetics Service, Laboratory Genetics, Aberdeen Royal Infirmary, Foresterhill, Aberdeen, AB25 2ZD, UK; Department of Obstetrics and Gynaecology, Faculty of Medical and Health Sciences, The University of Auckland, Auckland 1142, New Zealand; Centre for Cancer Research, Faculty of Medical and Health Sciences, The University of Auckland, Auckland 1142, New Zealand; Department of Ophthalmology, Faculty of Medical and Health Sciences, The University of Auckland, Auckland 1142, New Zealand; Department of Ophthalmology, Leuven University Hospital, Louvain, Belgium; Centre of Human Genetics, University Hospitals Leuven, Louvain, Belgium; Wessex Regional Genomics Laboratory, Salisbury District Hospital, Salisbury, Wiltshire, SP2 8BJ, UK; Faculty of Medicine, University of Southampton, Southampton, Hampshire, SO16 6YD, UK

## Abstract

Heterozygous *FOXL2* (non-)coding sequence and structural variants (SVs) lead to blepharophimosis, ptosis and epicanthus inversus syndrome (BPES), a rare, autosomal dominant developmental disorder characterized by a completely penetrant eyelid malformation and incompletely penetrant primary ovarian insufficiency (POI).

We collected variants from our in-house database, generated via clinical genetic testing and downstream research testing in the Center for Medical Genetics Ghent, Belgium (2001-2024), and via literature and other resources in the same period. All retrieved variants were categorized using ACMG/AMP classifications to increase the knowledge of pathogenicity.

We collected 413 unique genetic defects of the *FOXL2* region, including 76 novel variants, in 864 index patients. Of these, 87% of patients were identified with a coding *FOXL2* sequence variant. The polyalanine tract is a known mutational hotspot of *FOXL2*, illustrated here by the high percentage of pathogenic polyalanine expansions (24%). Furthermore, the molecular spectrum in typical BPES index patients is characterized by 8% coding deletions and 3% deletions located up- and downstream of *FOXL2*. The remaining 2% carry translocations along with chromosomal rearrangements of 3q23.

This uniform and structured reclassification, incorporating the largest dataset of variants implicated in *FOXL2*-associated disease so far, will improve both the diagnosis as well as genetic counselling for individuals with BPES.

## 1. Introduction

*FOXL2* is a 2.9 kb single-exon gene located on the long arm of chromosome 3 (3q22.3). The gene encodes a 376-amino-acid transcription factor with a highly conserved DNA-binding forkhead domain. In addition to this forkhead domain, a 14-amino-acid polyalanine tract can be distinguished, of which the function remains largely unknown (**Fig 1, A**) (1–3).

**Fig 1:**
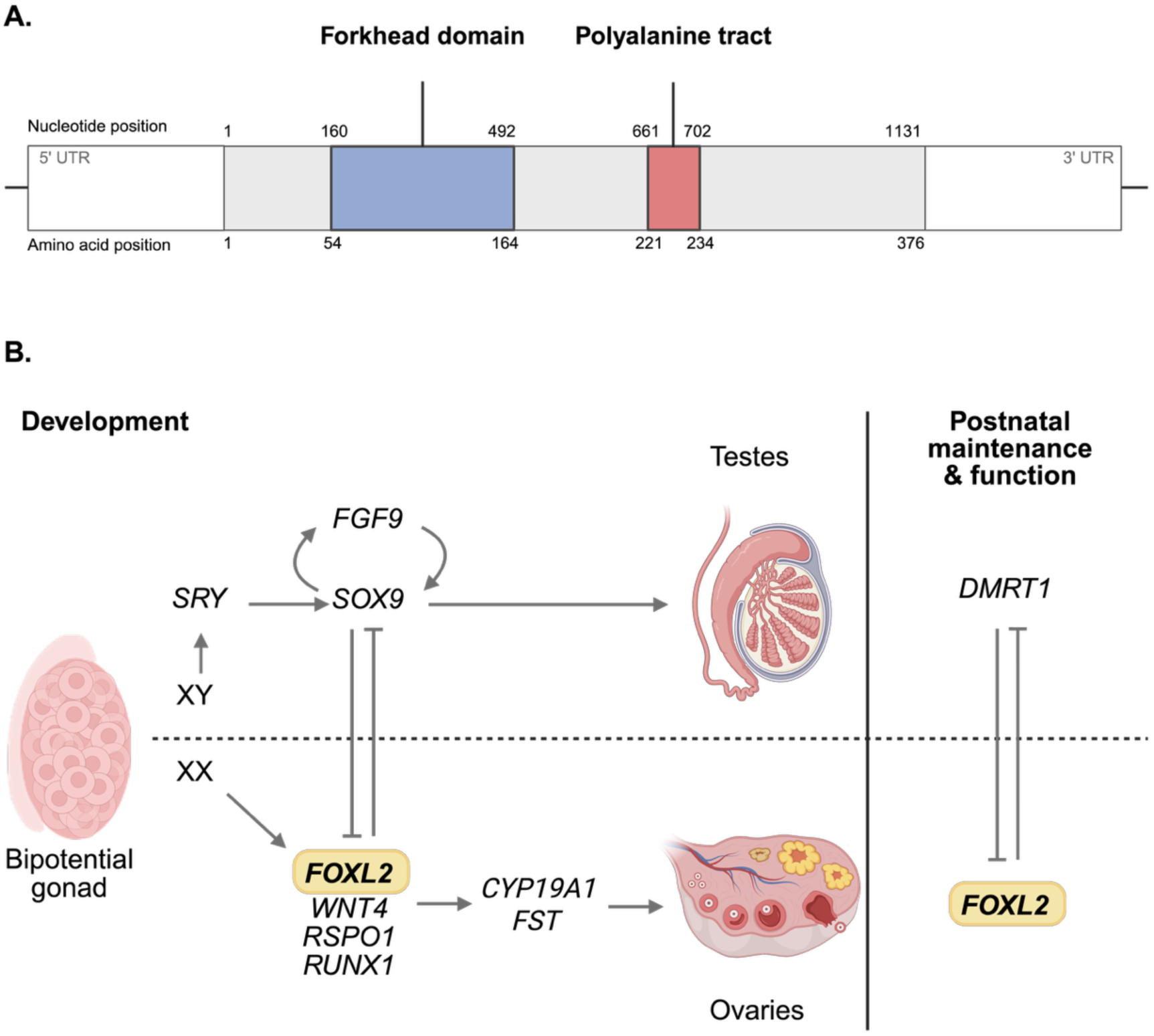
A) The FOXL2 protein comprises 376 amino acids and contains two key domains: the forkhead domain in blue (p.54–164) and the polyalanine tract in red (p.221–234). Adapted from Beysen *et al*., 2009 (17) B) The transcription factor FOXL2 is essential for ovarian development. At 5-6 weeks of gestation, the bipotential gonad differentiates toward either testis or ovaries, depending on the chromosomal sex. This process is tightly regulated by antagonistic interactions between key factors. FOXL2 inhibits SOX9, thereby repressing the pro-testis pathway and, in concert with other regulators, promoting ovarian differentiation. In adulthood, FOXL2 remains crucial for ovarian maintenance and function. Figure adapted from Tucker *et al.,* 2022 (6).

The FOXL2 transcription factor, like other members of the forkhead transcription factor family, is involved in key developmental processes and requires tightly controlled spatiotemporal gene regulation (4). During embryonic development, FOXL2 is expressed in the developing periocular mesenchyme, where it activates α smooth muscle actin (α-SMA), facilitating the formation of the levator smooth muscle (5). In developing ovaries, FOXL2 functions alongside other pro-female factors, promoting the upregulation of ovarian-specific genes while simultaneously suppressing testis-specific factors such as SOX9 (6–8) (**Fig 1, B**). Postnatally, *FOXL2* is expressed in the granulosa cells (i.e. supporting cells of the oocyte), pituitary gland, and uterus (6,9–12). In adult granulosa cells, FOXL2 is critical for ovarian maintenance and folliculogenesis, ensuring the integrity of ovarian function throughout reproductive life (13) (**Fig 1, B**). In line with other highly conserved transcription factor genes, impairment of *FOXL2* results in developmental disease and cancer.

Heterozygous germline variants in *FOXL2* are known to cause blepharophimosis, ptosis and epicanthus inversus syndrome (BPES; OMIM #110100), a rare, autosomal dominant, developmental disorder that can arise *de novo* or be inherited (2,3,14). BPES is characterized by a fully penetrant eyelid malformation and an incompletely penetrant and variable primary ovarian insufficiency (POI). The eyelid malformation is described according to four typical features including (1) horizontal (blepharophimosis) and (2) vertical (ptosis) narrowing of the palpebral fissures, (3) a skin fold running inwards and upwards from the lower eyelid (epicanthus inversus) and (4) a lateral displacement of the inner canthi (telecanthus). POI is defined as the absence of menses for over four months before the age of 40, resulting in early menopause and female subfertility or infertility due to impaired folliculogenesis in adult granulosa cells. BPES is associated with additional features including thick eyebrows, displacement of the lacrimal puncta, nasolacrimal drainage problems, a broad nasal bridge and low-set ears (10,14,15).

At the somatic level, *FOXL2* variants are strongly implicated in adult-type granulosa cell tumours, with a recurrent missense variant c.402C>G (p.Cys134Trp) detected in approximately 90-95% of cases (16).

In this study, we compiled a large collection of in-house variants identified through clinical and research genetic testing at our Center (Center for Medical Genetics Ghent, Ghent University Hospital, Ghent, Belgium) and elsewhere (Wessex Regional Genomics Laboratory, Salisbury District Hospital, Salisbury, UK; Institute of Human Genetics, University of Würzburg, Würzburg, Germany; Laboratory of Cytogenetics and Molecular Genetics, University Children’s Hospital, Krakow, Poland; Clinical Genetics Centre, Aberdeen Royal Infirmary, Aberdeen, UK; Centre of Human Genetics, University Hospitals Leuven, Louvain, Belgium) in index patients with a typical BPES phenotype. This dataset was complemented with previously published *FOXL2* sequence and structural variants (SVs) affecting the *FOXL2* region, all identified in patients with typical BPES. Together, this resulted in a comprehensive compendium of 413 distinct heterozygous constitutional *FOXL2* variants, ranging from coding sequence changes and copy number variants (CNVs) to translocations and non-coding CNVs disrupting putative *cis*-regulatory elements (CREs). All sequence variants were subsequently classified according to the most recent American College of Medical Genetics and Genomics and the Association for Molecular Pathology (ACMG-AMP) standard guidelines. Notably, this large study enabled us to expand the variant spectrum with 76 novel *FOXL2* genetic defects.

## 2. Material and methods

### 2.1 Literature search on patients with BPES and a confirmed FOXL2 genetic defect

We collected publications up to December 2024 that reported typical BPES patients with a genetically confirmed heterozygous *FOXL2* variant, using the following PubMed search terms: “*FOXL2*” [All Fields] AND (“mutation” [All Fields] OR “variant” [All Fields] OR “translocation” [All Fields] OR “deletion” [All Fields]). The literature search was extended via Google Scholar to not exclude publications that were not listed in PubMed. Moreover, we mined the ClinVar and Leiden Open-source Variation Database (LOVD) databases to include variants that were only submitted to the databases. Likewise, to ensure that unpublished SVs were not overlooked, the Genomics England and Decipher databases were consulted.

For all included variants, variant annotations, gnomAD allele frequencies (gnomAD v4.1.0), disease phenotype and patient information (including age and sex) were collected, if available. Duplicates were excluded from the variant collection based on overlap in patient information and author names of the included publications. In addition, studies for which the full text was unavailable or not accessible through the Ghent University Library, or not available in English, were excluded.

Although a comprehensive search was performed, we cannot exclude that certain *FOXL2* variants associated with BPES were missed due to the limitations of literature search tools (e.g. *FOXL2* variants solely mentioned in publications’ supporting information might have been overlooked).

### 2.2 Unreported patients with BPES and a confirmed molecular cause from the in-house patient cohort

In addition to variants obtained from literature and public databases, *FOXL2* variants identified through clinical genetic testing at the Center for Medical Genetics Ghent (Ghent University Hospital, Ghent, Belgium); Wessex Regional Genomics Laboratory (Salisbury District Hospital, Salisbury, UK); Institute of Human Genetics (University of Würzburg, Würzburg, Germany); Laboratory of Cytogenetics and Molecular Genetics (University Children’s Hospital, Krakow, Poland); Clinical Genetics Centre (Aberdeen Royal Infirmary, Aberdeen, UK) and Centre of Human Genetics (University Hospitals Leuven, Louvain, Belgium) were included. Given the specificity of the BPES phenotype and *FOXL2* being the only known disease locus for this rare syndrome, Sanger sequencing was performed across the *FOXL2* open reading frame in all cases with BPES. The initial polymerase chain reaction (PCR) to amplify the region of interest was performed using 3 overlapping primers pairs (**Table S4**) and Kapa2G Robust Master Mix (2x, Kapa Biosystems, MA, USA), followed by Sanger sequencing using the BrilliantDye^TM^ kit (v3.1, NimaGent, Nijmegen, the Netherlands) and ran on an ABI3730XL DNA analyzer (Applied Biosystems, MA, USA). Due to the GC-rich nature of the *FOXL2* region, 5% dimethyl sulfoxide (DMSO) was added to increase sequencing signal intensity.

Multiplex ligation-dependent probe amplification (MLPA) was performed using a commercially available probe mix (P054, MRC Holland, Amsterdam, the Netherlands) to detect CNVs. The MLPA kit contains 3 probes covering the coding *FOXL2* region, four probes covering the *ATR* region (exon 1; 4; 22 and 47), and five probes overlapping with the *PISRT1* region, a long non-coding RNA (lncRNA) located in the regulatory region upstream of *FOXL2*. Apart from deletion identification itself, it is also essential to size the deletion, which was previously done using targeted arrays and qPCR (18–20).

### 2.3 Nomenclature

Variant annotations in the *FOXL2* coding region were (re)assessed using Alamut (GRCh38) for coding DNA (c.), protein (p.), and genome (g.) nomenclature. When necessary, annotations were revised to align with the latest Human Genome Variation Society (HGVS) nomenclature guidelines (hgvs-nomenclature.org).

### 2.4 ACMG/AMP variant pathogenicity classification

All collected coding *FOXL2* variants were (re)classified using an in-house variant classification tool (VCT) (Center for Medical Genetics Ghent, Ghent University Hospital, Ghent, Belgium), developed in accordance with ACMG/AMP guidelines described by Richards S. *et al*. 2015 and Tavtigian *et al.*, 2018 (21,22). The VCT categorizes variants into five classes based on their calculated p-value, namely benign (p < 0.001), likely benign (0.001 ≤ p < 0.1), unknown significance (VUS) (0.1 ≤ p < 0.9), likely pathogenic (0.9 ≤ p < 0.99) or pathogenic (p ≥ 0.99).

### 2.5 LOVD and ClinVar submission

Following the (re)classification of all coding *FOXL2* sequence variants, they were uploaded to the LOVD and ClinVar database to ensure comprehensive documentation of all collected *FOXL2* variants. When available, patient information and *in silico* predictions were also included.

This work was conducted according to the tenets of the Declaration of Helsinki and was approved by the data access committee of Ghent University Hospital.

## 3. Results

### 3.1 Collection of BPES index cases and unique FOXL2 genetic defects

Through a comprehensive literature and database search, we collected 497 index cases with a typical BPES phenotype that have been reported previously in international, peer-reviewed literature and curated databases, including ClinVar, Genomics England and DECIPHER (2,3,14,18–20,23–117). In addition, we report 367 BPES index patients with a genetic defect of the *FOXL2* region identified via routine diagnostic pipelines, resulting in a total collection of 864 index cases included in this study (**Table 1, S1, S2 & S3**). This large BPES patient cohort revealed 413 unique *FOXL2* genetic defects, both in the coding and non-coding regions of *FOXL2*. Notably, we report 76 novel variants associated with a typical BPES phenotype in this compendium (**Table 1**).

**Table 1:**
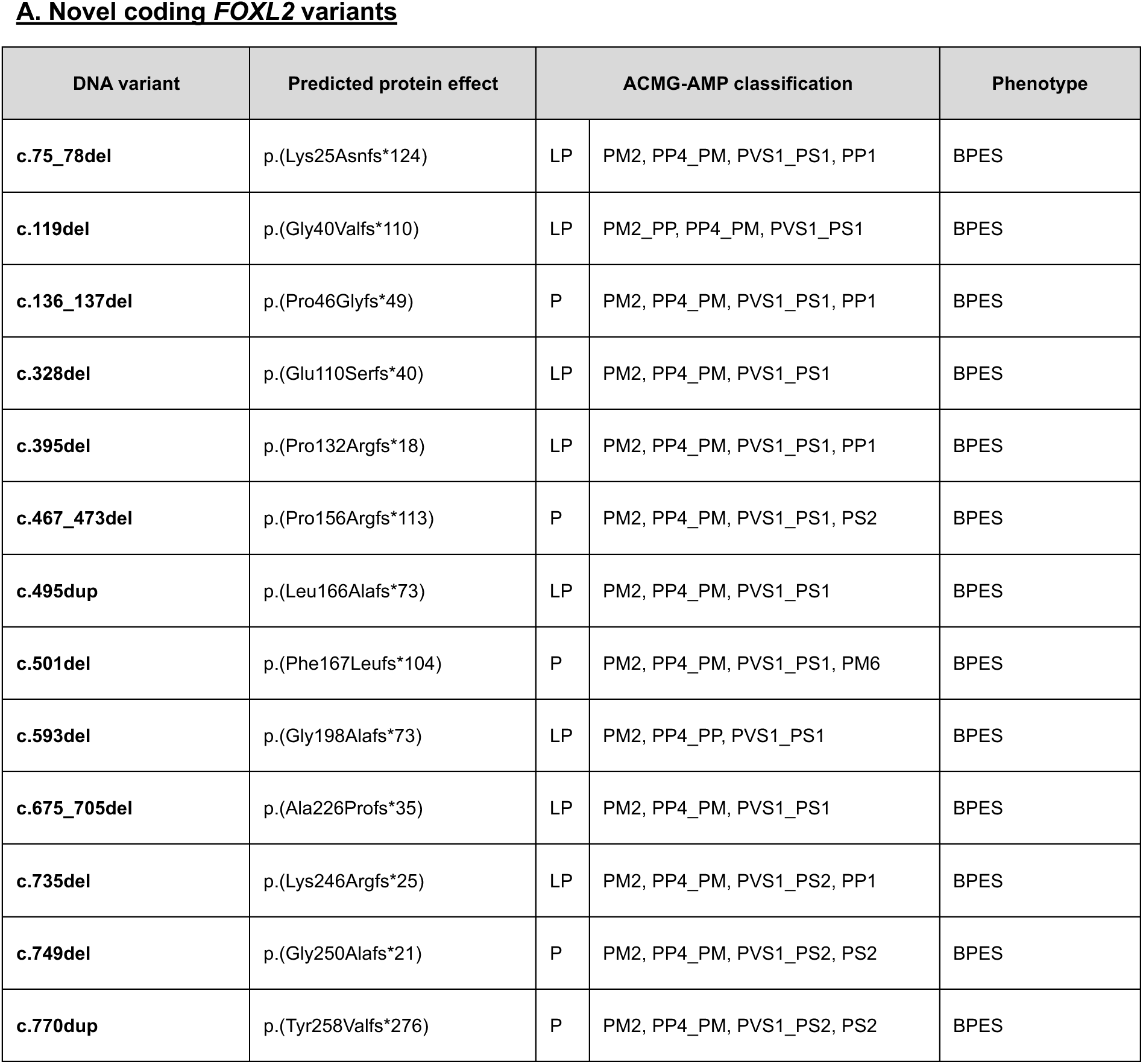
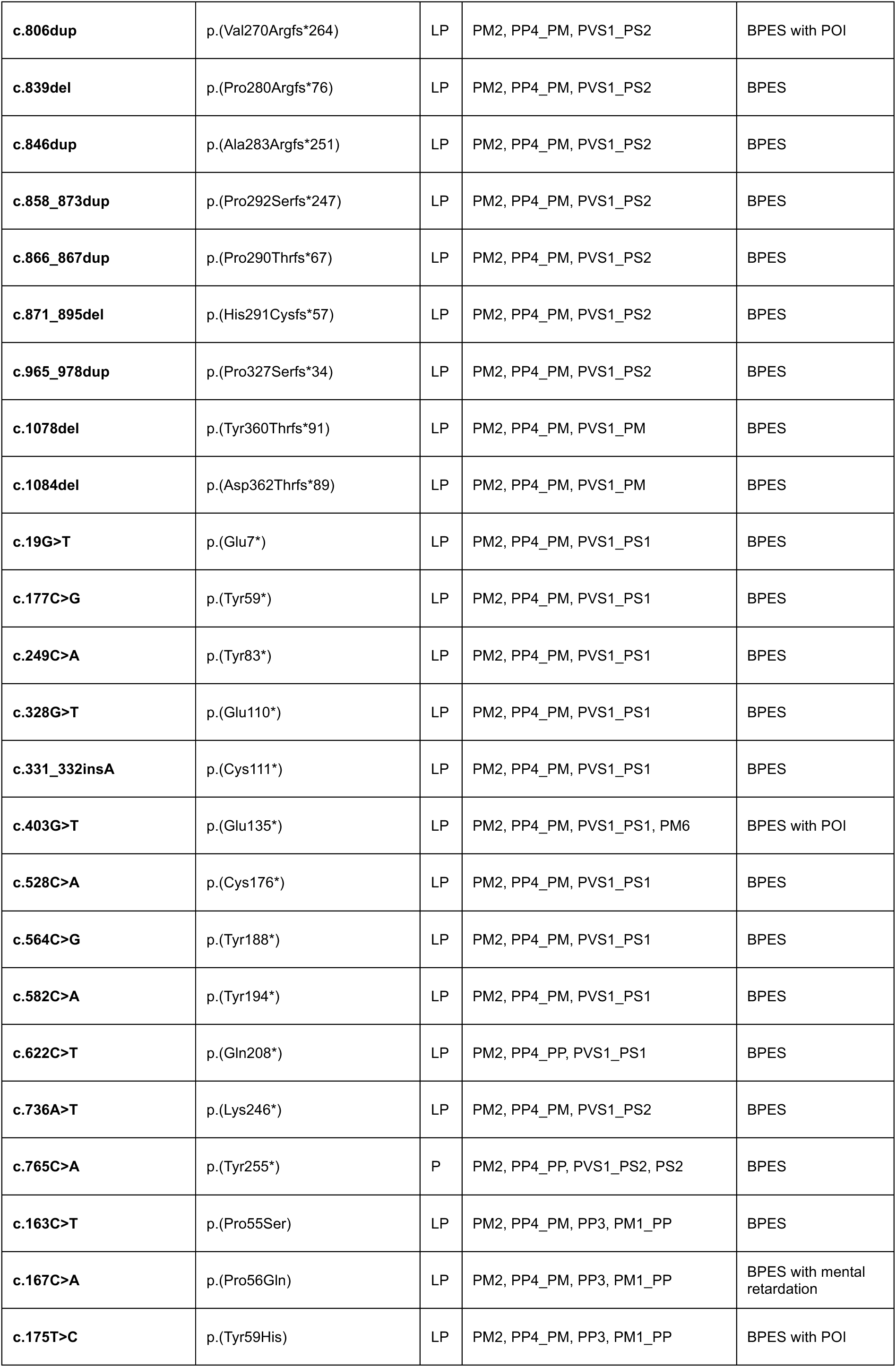
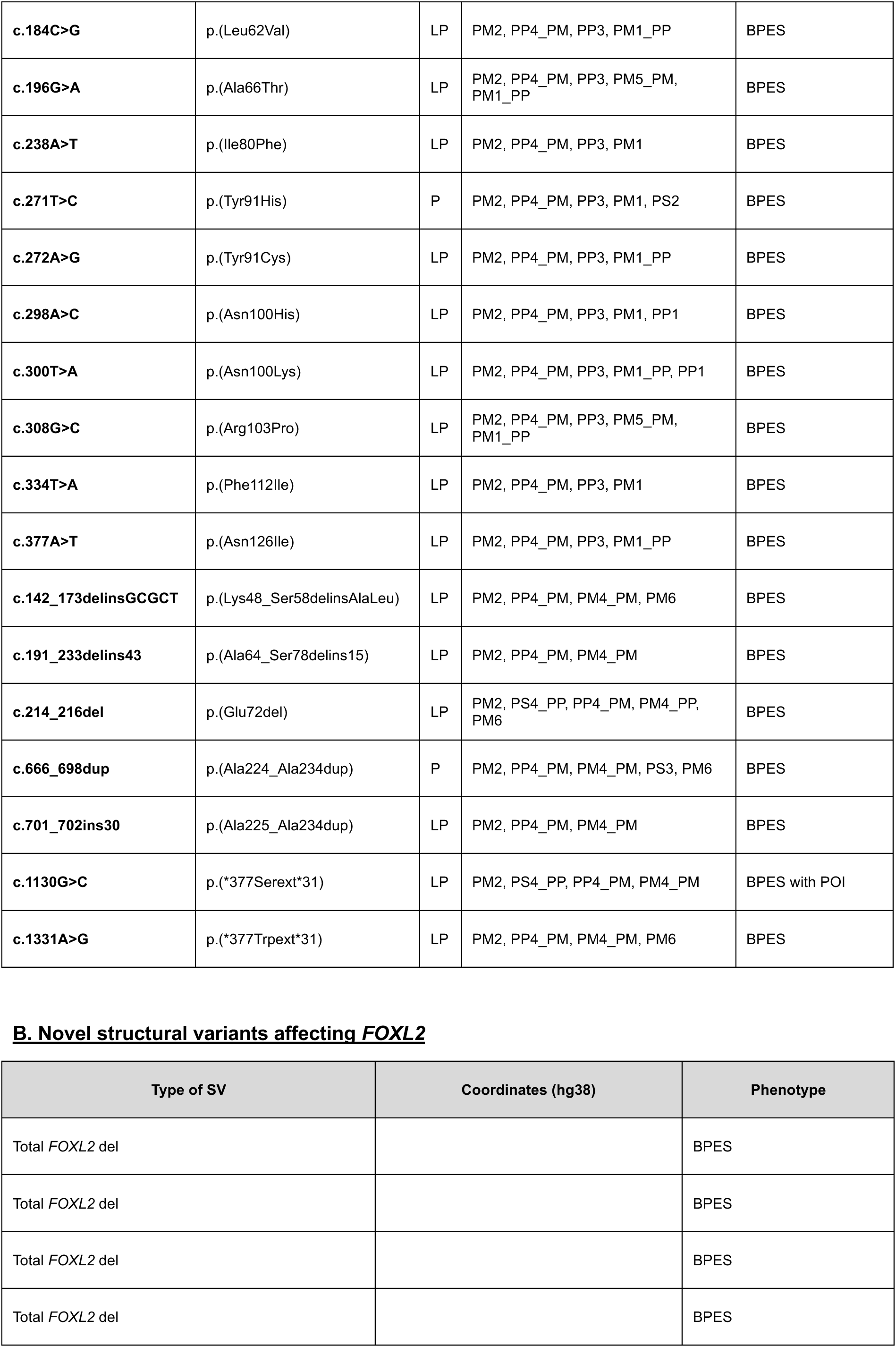
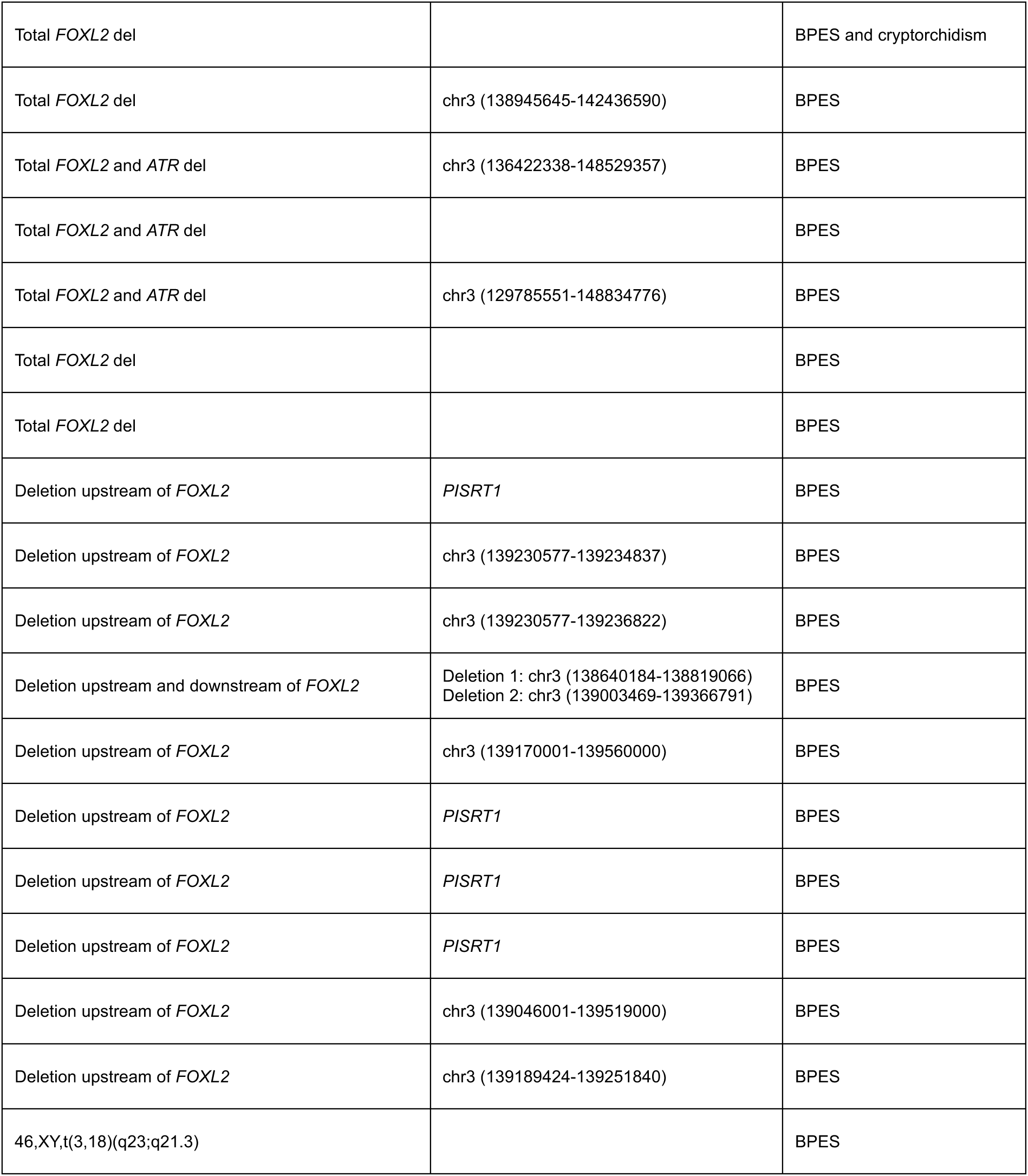
(A) We identified 54 novel *FOXL2* coding variants, including missense, nonsense, frameshift, and in-frame indels, in this study. All variants are annotated with their corresponding ACMG/AMP variant classifications. (B) We identified 22 SVs affecting the *FOXL2* locus in this study, including 11 deletions encompassing the entire *FOXL2* gene, 10 deletions located upstream of *FOXL2*, one combined upstream and downstream deletion flanking *FOXL2*, and one novel chromosomal translocation involving the *FOXL2* genomic region.

**A. Novel coding FOXL2 variants**
| DNA variant | Predicted protein effect | ACMG-AMP classification |  | Phenotype |
| --- | --- | --- | --- | --- |
| **c.75_78del** | p.(Lys25Asnfs*124) | LP | PM2, PP4_PM, PVS1_PS1, PP1 | BPES |
| **c.119del** | p.(Gly40Valfs*110) | LP | PM2_PP, PP4_PM, PVS1_PS1 | BPES |
| **c.136_137del** | p.(Pro46Glyfs*49) | P | PM2, PP4_PM, PVS1_PS1, PP1 | BPES |
| **c.328del** | p.(Glu110Serfs*40) | LP | PM2, PP4_PM, PVS1_PS1 | BPES |
| **c.395del** | p.(Pro132Argfs*18) | LP | PM2, PP4_PM, PVS1_PS1, PP1 | BPES |
| **c.467_473del** | p.(Pro156Argfs*113) | P | PM2, PP4_PM, PVS1_PS1, PS2 | BPES |
| **c.495dup** | p.(Leu166Alafs*73) | LP | PM2, PP4_PM, PVS1_PS1 | BPES |
| **c.501del** | p.(Phe167Leufs*104) | P | PM2, PP4_PM, PVS1_PS1, PM6 | BPES |
| **c.593del** | p.(Gly198Alafs*73) | LP | PM2, PP4_PP, PVS1_PS1 | BPES |
| **c.675_705del** | p.(Ala226Profs*35) | LP | PM2, PP4_PM, PVS1_PS1 | BPES |
| **c.735del** | p.(Lys246Argfs*25) | LP | PM2, PP4_PM, PVS1_PS2, PP1 | BPES |
| **c.749del** | p.(Gly250Alafs*21) | P | PM2, PP4_PM, PVS1_PS2, PS2 | BPES |
| **c.770dup** | p.(Tyr258Valfs*276) | P | PM2, PP4_PM, PVS1_PS2, PS2 | BPES |
| **c.806dup** | p.(Val270Argfs*264) | LP | PM2, PP4_PM, PVS1_PS2 | BPES with POI |
| **c.839del** | p.(Pro280Argfs*76) | LP | PM2, PP4_PM, PVS1_PS2 | BPES |
| **c.846dup** | p.(Ala283Argfs*251) | LP | PM2, PP4_PM, PVS1_PS2 | BPES |
| **c.858_873dup** | p.(Pro292Serfs*247) | LP | PM2, PP4_PM, PVS1_PS2 | BPES |
| **c.866_867dup** | p.(Pro290Thrfs*67) | LP | PM2, PP4_PM, PVS1_PS2 | BPES |
| **c.871_895del** | p.(His291Cysfs*57) | LP | PM2, PP4_PM, PVS1_PS2 | BPES |
| **c.965_978dup** | p.(Pro327Serfs*34) | LP | PM2, PP4_PM, PVS1_PS2 | BPES |
| **c.1078del** | p.(Tyr360Thrfs*91) | LP | PM2, PP4_PM, PVS1_PM | BPES |
| **c.1084del** | p.(Asp362Thrfs*89) | LP | PM2, PP4_PM, PVS1_PM | BPES |
| **c.19G>T** | p.(Glu7*) | LP | PM2, PP4_PM, PVS1_PS1 | BPES |
| **c.177C>G** | p.(Tyr59*) | LP | PM2, PP4_PM, PVS1_PS1 | BPES |
| **c.249C>A** | p.(Tyr83*) | LP | PM2, PP4_PM, PVS1_PS1 | BPES |
| **c.328G>T** | p.(Glu110*) | LP | PM2, PP4_PM, PVS1_PS1 | BPES |
| **c.331_332insA** | p.(Cys111*) | LP | PM2, PP4_PM, PVS1_PS1 | BPES |
| **c.403G>T** | p.(Glu135*) | LP | PM2, PP4_PM, PVS1_PS1, PM6 | BPES with POI |
| **c.528C>A** | p.(Cys176*) | LP | PM2, PP4_PM, PVS1_PS1 | BPES |
| **c.564C>G** | p.(Tyr188*) | LP | PM2, PP4_PM, PVS1_PS1 | BPES |
| **c.582C>A** | p.(Tyr194*) | LP | PM2, PP4_PM, PVS1_PS1 | BPES |
| **c.622C>T** | p.(Gln208*) | LP | PM2, PP4_PP, PVS1_PS1 | BPES |
| **c.736A>T** | p.(Lys246*) | LP | PM2, PP4_PM, PVS1_PS2 | BPES |
| **c.765C>A** | p.(Tyr255*) | P | PM2, PP4_PP, PVS1_PS2, PS2 | BPES |
| **c.163C>T** | p.(Pro55Ser) | LP | PM2, PP4_PM, PP3, PM1_PP | BPES |
| **c.167C>A** | p.(Pro56Gln) | LP | PM2, PP4_PM, PP3, PM1_PP | BPES with mental retardation |
| **c.175T>C** | p.(Tyr59His) | LP | PM2, PP4_PM, PP3, PM1_PP | BPES with POI |
| **c.184C>G** | p.(Leu62Val) | LP | PM2, PP4_PM, PP3, PM1_PP | BPES |
| **c.196G>A** | p.(Ala66Thr) | LP | PM2, PP4_PM, PP3, PM5_PM, PM1_PP | BPES |
| **c.238A>T** | p.(Ile80Phe) | LP | PM2, PP4_PM, PP3, PM1 | BPES |
| **c.271T>C** | p.(Tyr91His) | P | PM2, PP4_PM, PP3, PM1, PS2 | BPES |
| **c.272A>G** | p.(Tyr91Cys) | LP | PM2, PP4_PM, PP3, PM1_PP | BPES |
| **c.298A>C** | p.(Asn100His) | LP | PM2, PP4_PM, PP3, PM1, PP1 | BPES |
| **c.300T>A** | p.(Asn100Lys) | LP | PM2, PP4_PM, PP3, PM1_PP, PP1 | BPES |
| **c.308G>C** | p.(Arg103Pro) | LP | PM2, PP4_PM, PP3, PM5_PM, PM1_PP | BPES |
| **c.334T>A** | p.(Phe112Ile) | LP | PM2, PP4_PM, PP3, PM1 | BPES |
| **c.377A>T** | p.(Asn126Ile) | LP | PM2, PP4_PM, PP3, PM1_PP | BPES |
| **c.142_173delinsGCGCT** | p.(Lys48_Ser58delinsAlaLeu) | LP | PM2, PP4_PM, PM4_PM, PM6 | BPES |
| **c.191_233delins43** | p.(Ala64_Ser78delins15) | LP | PM2, PP4_PM, PM4_PM | BPES |
| **c.214_216del** | p.(Glu72del) | LP | PM2, PS4_PP, PP4_PM, PM4_PP, PM6 | BPES |
| **c.666_698dup** | p.(Ala224_Ala234dup) | P | PM2, PP4_PM, PM4_PM, PS3, PM6 | BPES |
| **c.701_702ins30** | p.(Ala225_Ala234dup) | LP | PM2, PP4_PM, PM4_PM | BPES |
| **c.1130G>C** | p.(*377Serext*31) | LP | PM2, PS4_PP, PP4_PM, PM4_PM | BPES with POI |
| **c.1331A>G** | p.(*377Trpext*31) | LP | PM2, PP4_PM, PM4_PM, PM6 | BPES |

**Table 1:** (A) We identified 54 novel *FOXL2* coding variants, including missense, nonsense, frameshift, and in-frame indels, in this study.
| Type of SV | Coordinates (hg38) | Phenotype |
| --- | --- | --- |
| Total *FOXL2* del |  | BPES |
| Total *FOXL2* del |  | BPES |
| Total *FOXL2* del |  | BPES |
| Total *FOXL2* del |  | BPES |
| Total *FOXL2* del |  | BPES and cryptorchidism |
| Total *FOXL2* del | chr3 (138945645-142436590) | BPES |
| Total *FOXL2* and *ATR* del | chr3 (136422338-148529357) | BPES |
| Total *FOXL2* and *ATR* del |  | BPES |
| Total *FOXL2* and *ATR* del | chr3 (129785551-148834776) | BPES |
| Total *FOXL2* del |  | BPES |
| Total *FOXL2* del |  | BPES |
| Deletion upstream of *FOXL2* | *PISRT1* | BPES |
| Deletion upstream of *FOXL2* | chr3 (139230577-139234837) | BPES |
| Deletion upstream of *FOXL2* | chr3 (139230577-139236822) | BPES |
| Deletion upstream and downstream of *FOXL2* | Deletion 1: chr3 (138640184-138819066)<br>Deletion 2: chr3 (139003469-139366791) | BPES |
| Deletion upstream of *FOXL2* | chr3 (139170001-139560000) | BPES |
| Deletion upstream of *FOXL2* | *PISRT1* | BPES |
| Deletion upstream of *FOXL2* | *PISRT1* | BPES |
| Deletion upstream of *FOXL2* | *PISRT1* | BPES |
| Deletion upstream of *FOXL2* | chr3 (139046001-139519000) | BPES |
| Deletion upstream of *FOXL2* | chr3 (139189424-139251840) | BPES |
| 46,XY,t(3,18)(q23;q21.3) |  | BPES |

### 3.2 FOXL2 coding sequence variants and their variant classification

A total of 752 index patients has been identified with a *FOXL2* coding sequence variant making up 87% (752/864) of the total solved BPES index cases and revealed 54 unique novel variants in the coding region of the *FOXL2* gene. The majority of these index patients are heterozygous for a frameshift variant (39%; 295/752), followed by in-frame indels (31%; 231/752), missense variants (18%; 137/752), nonsense variants (11%; 82/752) and stop loss variants (1%; 7/752) (**Table S1, Fig 2**).

**Fig 2:**
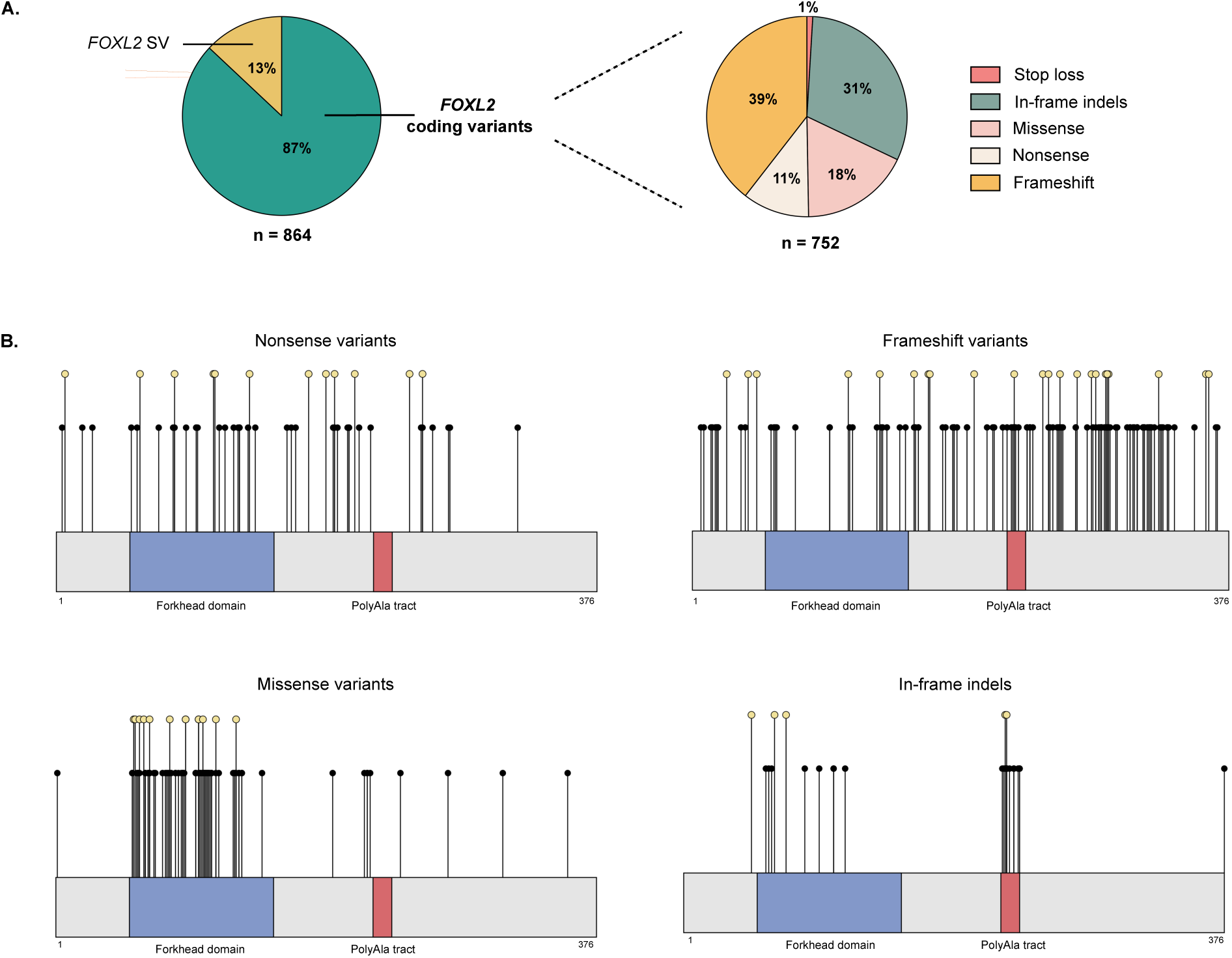
A) In 87% of the BPES patient cohort (n = 752), variants were identified in the coding region of *FOXL2*. These include frameshift (39%), in-frame indel (31%), missense (18%), nonsense (11%), and stop-loss (1%) variants. B) Coding *FOXL2* variants are distributed across the entire protein. Missense variants cluster in the forkhead domain, while in-frame indels predominantly affect the polyalanine tract, both known mutational hotspots of the *FOXL2* region. Novel variants identified in this study are highlighted in yellow.

#### Frameshift variants

Our compendium contains 135 unique frameshift variants, including 22 novel variants, of which 24% (32/135) are classified as pathogenic and 76% (103/134) are classified as likely pathogenic. Frameshifts of the *FOXL2* coding region result in either a predicted truncated protein (100/135) or an elongated protein (35/135), leading to loss-of-function. Notably, the most recurrent frameshift variants, including c.843_859dup (p.[Pro287Argfs*75]) (n=66), c.855_871dup (p.[His291Argfs*71]) (n=38) and c.804dup (p.[Gly269Argfs*265]) (n=20), are clustered downstream of the polyalanine tract (**Table S1, Fig 2**).

#### In-frame indels

In addition to the 23 previously reported unique in-frame indel variants, we report 3 novel in-frame variants located outside the polyalanine tract (c.142_173delinsGCGCT, p.[Lys48_Ser58delinsAlaLeu]; c.191_233delins43, p.[Ala64_Ser78delins15] and c.214_216del, p.[Glu72del]), and 2 novel in-frame duplications located within the polyalanine tract (c.666_698dup, p.[Ala224_Ala234dup] and c.701_702ins30, p.[Ala225_Ala234dup]) in this study. Of all unique variants, 32% are classified as pathogenic (9/28) and 68% as likely pathogenic (19/28).

To date, 12 unique duplications, and one deletion in the polyalanine region have been described. Polyalanine expansions are the most frequently reported type of in-frame indels, and to a larger extent, the most frequently reported *FOXL2* variants in BPES, as they are identified in 209 BPES index patients in this large cohort (24 %; 209/864). The most prevalent polyalanine expansion is c.672_701(p.[Ala225_Ala234dup]), observed in at least 156 BPES index cases, including 69 from our patient cohort. Another common polyalanine expansion, c.663_692dup (p.[Ala225_Ala234dup]), has been reported 13 times in literature and is also found in 21 index cases within our patient cohort (**Table S1, Fig 2 & 3**).

**Fig 3:**
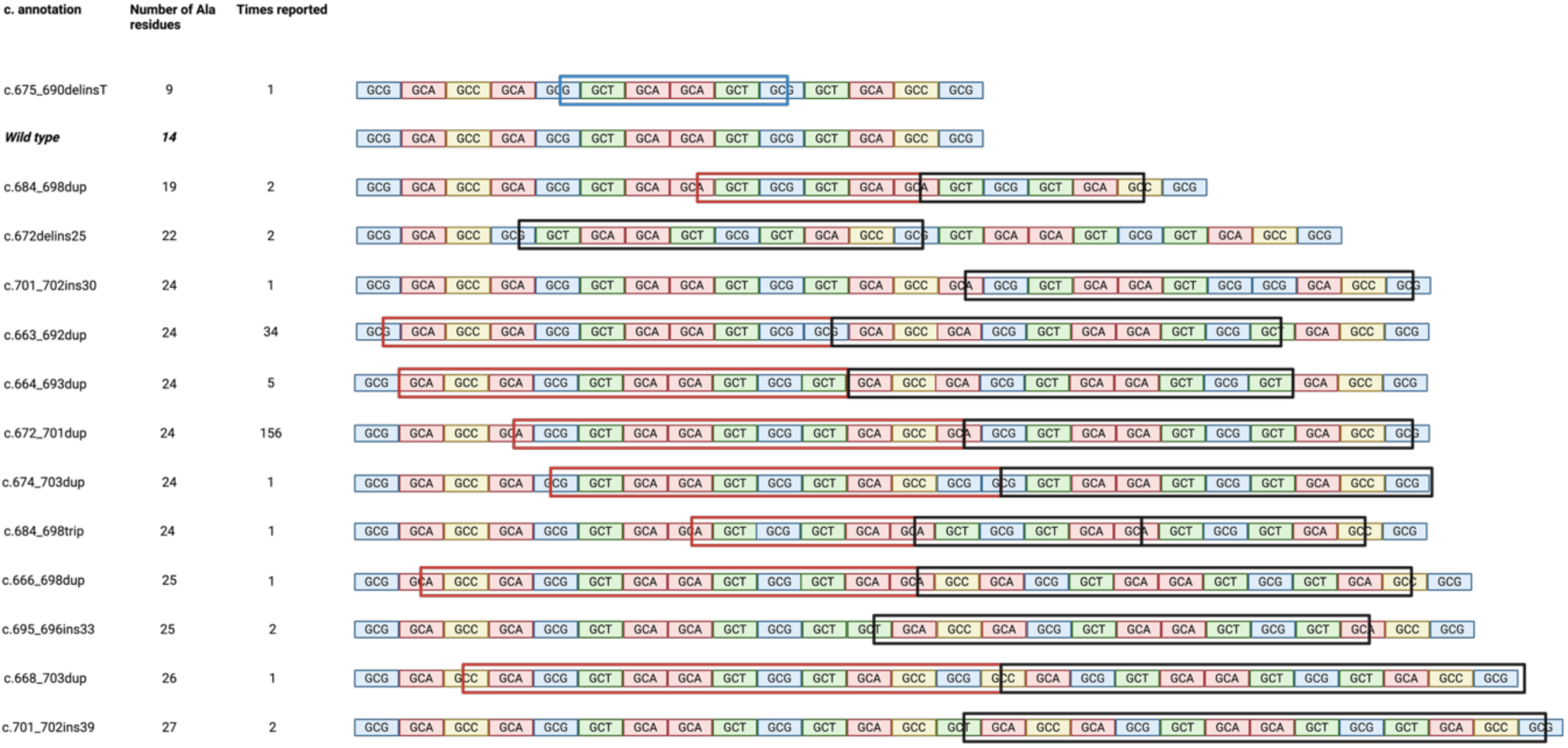
Polyalanine expansions were identified in 24% of the BPES patient cohort, making this the most frequent variant type. In most cases, the polyalanine tract is expanded from 14 to 24 alanine residues. Additionally, one contraction of the polyalanine tract was observed. The blue bar indicates deleted amino acids, the red bar represents the original repeat that was duplicated, and the black bar denotes the inserted repeat.

Interestingly, a small polyalanine expansion of 5+ alanine residues (c.684_698dup; p.[Ala230_Ala234dup]) was reported in an Indian, multigenerational family. Heterozygous family members remained (seemingly) unaffected, whereas homozygous individuals had a typical BPES phenotype. One of the homozygous female family members was also diagnosed with POI (78). Notably, the same variant was identified in an unrelated, sporadic case in a heterozygous state with a typical BPES phenotype (104) (**Table S1**).

#### Missense variants

Next, 84 unique missense variants were found of which the majority are classified as pathogenic (33%; 28/84) or likely pathogenic (61%; 51/84), and 6% as VUS (5/84). Most (n=74) are in located the forkhead domain, which is known to be a mutational hotspot for missense variants in *FOXL2*. We have identified 13 novel missense variants; all located within the forkhead domain (**Table S1, Fig 2**).

Single nucleotide polymorphisms (SNPs) have previously been reported in the *FOXL2* region, including c.501C>T (p.Phe167=), c.536C>G (p.Ala179Gly), c.647C>T (p.Ala216Val), and c.869C>A (p.Pro290His) (60,64,74,90,118). These variants have occasionally been detected in patients with BPES; however, their presence has also been documented in unaffected individuals, and they are catalogued in public SNP databases (e.g.; dbSNP: rs7432551, rs61750361, rs565208053). This suggests that they represent population polymorphisms rather than pathogenic variants. According to ACMG classification criteria, their relatively high allele frequencies in gnomAD, together with consistent *in silico* predictions of benign impact, further support that these variants should be considered (likely) benign.

#### Nonsense variants

We identified 51 unique nonsense variants in a total of 82 index patients, of which 12 are novel. All variants were classified as pathogenic (35%; 18/51) or likely pathogenic (65%; 33/51). Most of these variants are observed only once, with the notable exception of the recurrent variant c.655C>T (p.[Gln219*]), reported 10 times. Another nonsense variant, p.(Trp128*), has been reported 6 times, albeit resulting from different nucleotide changes (c.383G>A, c.384G>A, and c.384del) (**Table S1, Fig 2**).

#### Stop loss variants

Finally, a total of 3 unique stop-loss variants, all classified as likely pathogenic, have been identified in 7 BPES index cases. Among these, 2 are novel stop-loss variants, c.1130G>C (p.(*377Serext*31)) and c. 1331A>G (p.(*377Trpext*31)), found in 2 and 1 BPES index cases, respectively (**Table S1, Fig 2**).

### 3.3 Structural variants affecting the FOXL2 region

#### Copy number variants of the FOXL2 region

CNVs affecting the *FOXL2* gene were identified in 8% (71/864) of this BPES patient cohort and were detected using MLPA *FOXL2*-specific probes (**Table S2, Fig 4**). All but one of these CNVs encompassed the entire *FOXL2* gene. A single case involved a partial *FOXL2* deletion (BPES_10). The *ATR* gene, also covered by the MLPA assay, was co-deleted in approximately 23% of all detected deletions. Notably, all reported CNVs exclusively comprised deletions, with no instances of duplications associated with BPES documented to date. Furthermore, literature review revealed an additional 23 cases with *FOXL2* whole gene deletions, albeit lacking specific genomic coordinates. Via DECIPHER, we were able to identify 6 more patients with BPES harbouring a *FOXL2* (and *ATR*) deletion, with detailed genomic coordinates provided. In-house, 31 previously reported cases with BPES were molecularly diagnosed using MLPA followed by qPCR and/or microarray-based copy number analysis, enabling precise delineation of the deletions (18–20,118). Among the deletions that were further delineated, nearly all extended beyond *FOXL2* to encompass one or more neighbouring genes. Additionally, we report 11 novel deletions spanning the entire *FOXL2* region in this study.

**Fig 4:**
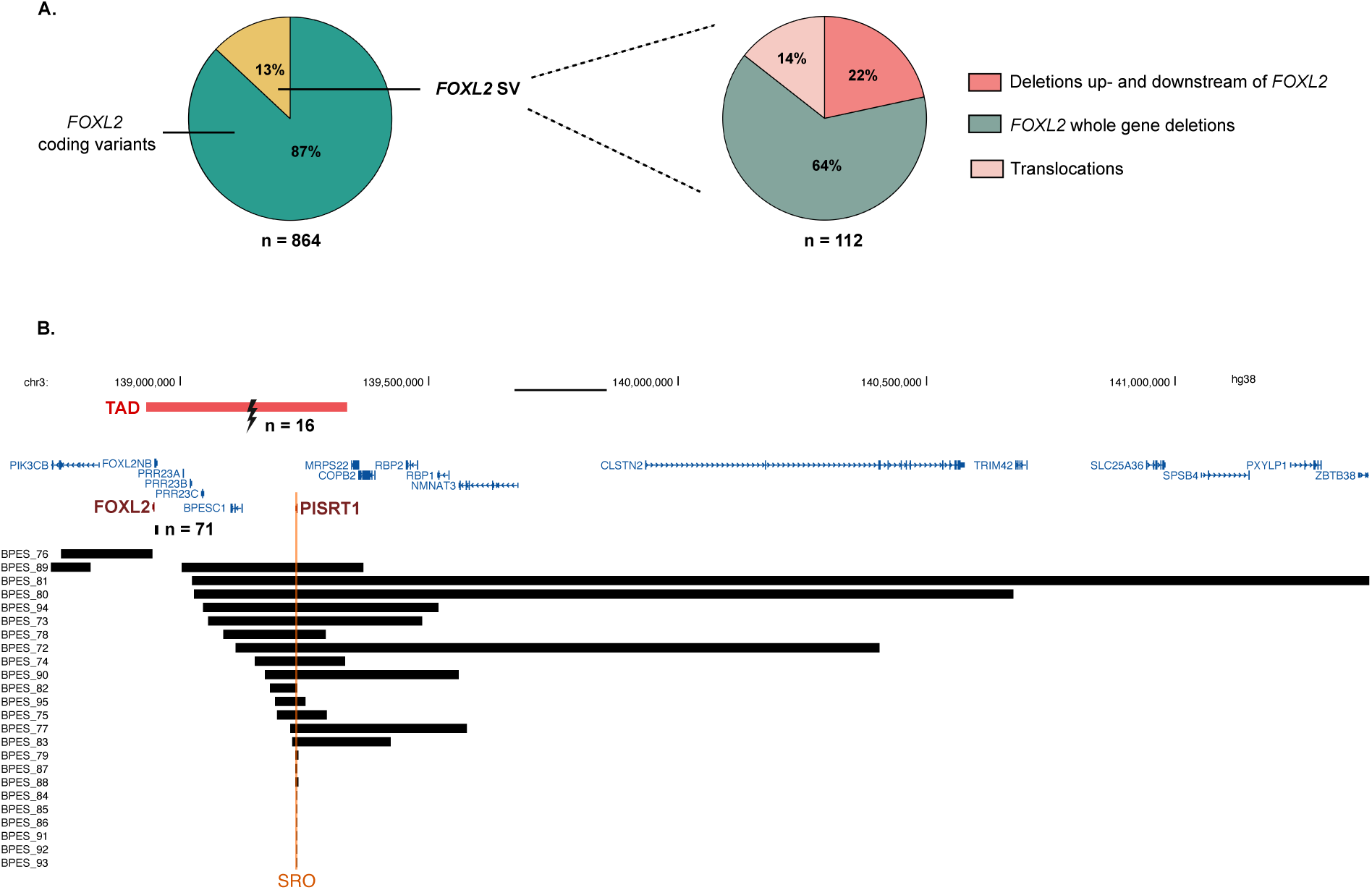
A) SVs were identified in 13% of the BPES patient cohort (n = 112). These include whole-gene deletions of *FOXL2* (64%), deletions located up-or downstream of *FOXL2* (22%), and translocations (14%). B) Translocation breakpoints cluster upstream of *FOXL2*, disrupting the regulatory landscape and physically separating the gene from its regulatory elements. Moreover, upstream deletions share an SRO (indicated in orange) of 4.5 kb, encompassing putative *FOXL2* enhancers, including the lncRNA *PISRT1*. The topologically associated domain (TAD) of *FOXL2* is highlighted with a red bar, marking the boundaries of its regulatory domain. Deletions for which no further delineation was performed; bars indicate whether a *FOXL2* or *PISRT1* deletion was detected by MLPA, although the exact genomic coordinates of these deletions remain undetermined.

Deletions disrupting the *FOXL2 cis*-regulatory region, including the lncRNA *PISRT1*, were identified in 3% (24/864) of genetically solved BPES index patients, ultimately leading to aberrant *FOXL2* expression (**Table S2, Fig 4**). In-house, 10 typical patients with BPES harbouring a *cis*-regulatory deletion could be identified, as previously published by us (18–20,118). One of the deletions is located downstream of *FOXL2*, whereas the other 9 deletions are located upstream of *FOXL2*. In (Bouman *et al.,* 2018), a non-coding 54-kb deletion (234 kb upstream of *FOXL2*) is described that encompasses the upstream *cis*-regulatory region including *PISRT1* in an infant female patient and her father, both displaying a typical BPES phenotype. Based on this regulatory deletion, the shortest region of overlap (SRO) of all non-coding deletions, located upstream of *FOXL2*, could be delineated to a region of 4573 bp (chr3: 139230309-139234882 [Hg38]) (35). Additionally, we describe 10 cases with BPES in this study with novel deletions upstream of *FOXL2*, all of which include the *PISRT1* region.

Importantly, one exceptional case (BPES_89) initially appeared to involve only a *PISRT1* deletion, as revealed by MLPA. However, subsequent CNV-sequencing analysis revealed the presence of two distinct deletions, located up- and downstream of *FOXL2*, indicating a more complex structural rearrangement of the *FOXL2* locus (**Table S2, Fig 4**).

Another interesting case described a Chinese family with BPES in which all affected males additionally exhibited polydactyly (BPES_96, **Table S2**). The causative variant was a deletion within the *FOXL2* promoter overlapping a CRE, and functional luciferase assays demonstrated reduced promoter activity, supporting a regulatory loss-of-function mechanism (119).

### Translocations

Lastly, cytogenetic rearrangements of 3q23 were found in 2% (16/864) of typical BPES index patients. Fifteen translocations had been described in literature before (2,94,106–111,113–115,120–124) (**Table S3, Fig 4**). Additionally, we identified one novel *de novo* translocation t(3,18)(q23;q21.3) in a male patient with BPES. None of the identified translocations disrupted the *FOXL2* coding sequence itself. Instead, all mapped breakpoints clustered upstream of *FOXL2* within its *cis*-regulatory domain, consistent with a position-effect mechanism in which disruption of long-range enhancers leads to *FOXL2* dysregulation, as previously shown (94,109,112,113,115).

## 4. Discussion

### Biological relevance

FOXL2 is a crucial transcription factor in both eyelid and ovarian development, but also postnatally FOXL2 remains critical for maintenance of the ovaries. Pathogenic variants in the *FOXL2* gene underlie BPES, associating an eyelid malformation with POI.

In this study, we compiled a comprehensive compendium of (virtually) all reported *FOXL2* genetic defects associated with BPES and of our in-house patient cohort, encompassing both (non-)coding sequence variants and SVs. Across a total of 864 index cases, we distinguished 413 unique genetic defects of the *FOXL2* region, including 76 novel variants.

The majority of *FOXL2* genetic defects are coding sequence variants, as they are identified in 87% of molecularly solved index patients with BPES.

As *FOXL2* is a single-exon gene, aberrant transcripts are expected to escape nonsense-mediated decay (NMD). Consequently, null variants introducing premature stop codons, are predicted to give rise to truncated FOXL2 proteins, missing the forkhead domain and/or the polyalanine tract (118). In contrast to the wild-type FOXL2, which displays exclusive nuclear localization, truncated FOXL2 proteins tend to form nuclear aggregates (26,125,126).

Missense variants represent 18% of all sequence variants and are predominantly clustered within the forkhead domain, a well-established mutational hotspot (**Fig 2**). Alterations in this domain are compromising DNA binding and transcriptional regulation, a mechanism that has been extensively studied previously (30,126–129).

A second mutational hotspot within the *FOXL2* gene is the polyalanine tract. In 24% of all BPES index patients, an expansion of the polyalanine tract is found, making this the most recurrent *FOXL2* variant associated with typical BPES. The WT polyalanine tract is located between position p.221 and p.234, consists of 14 alanine-residues and is strictly conserved amongst mammals (128,130). However, its function remains largely unknown. Up to date, 12 unique duplications, and one deletion of the polyalanine tract have been reported (**Table S1, Fig 3**), demonstrating that BPES can be caused by both expansion and contraction of the polyalanine tract, encoded by imperfect trinucleotide repeats. These expansions are most likely caused by slippage of DNA polymerase during replication of these trinucleotide repeats. It was shown previously that these expansions result in the misfolding of the protein and as such impair FOXL2 protein function in a length-dependent manner, resulting in protein mislocalization from the nucleus to the cytoplasm with cytoplasmic aggregation of the altered FOXL2 protein (130).

The remainder of *FOXL2* genetic defects consisted of deletions affecting the gene (8%) or its *cis*-regulatory region (3%), as well as upstream translocations separating *FOXL2* from its *cis*-regulatory domain (2%). These SVs are also expected to result in *FOXL2* loss-of-function through reduced or absent gene expression.

Interestingly, these non-coding SVs represent unique natural human models for studying the *cis*-regulation of *FOXL2*. We have previously delineated the shortest region of overlap (SRO) of regulatory deletions to a 7.4 kb region upstream of *FOXL2* (20). Recently, a novel deletion has been reported that delineates the SRO even further to a region of 4.5 kb (35), comprising a cluster of candidate CREs and a lncRNA *PISRT1*. Interestingly, the SRO delineated in human cases with BPES has its counterpart in a natural animal model, namely the Polled Intersex Syndrome (PIS) goat (131–133). First, the syntenic region affected by a complex non-coding SV in the Polled Intersex goat overlaps with the 4.5 kb SRO delineated in human BPES. Second, the Polled Intersex phenotype is characterized by the dominant absence of horns in both sexes, and recessive sex reversal in XX animals. Another animal model for aberrant *cis*-regulation of *FOXL2* is the piggyBac mouse (*Foxl2^PB/PB^*), caused by an insertion 160 kb upstream of *Foxl2*, leading to reduced *Foxl2* expression and causing an eyelid phenotype mimicking BPES (134). Although the syntenic regions in human, goat and mouse are not ultraconserved, the occurrence of cross-species non-coding defects leading to similar phenotypes supports the hypothesis of a functional conservation of the upstream *cis*-regulatory region of *FOXL2*. However, the precise function of this *cis*-regulatory region has remained elusive so far and requires further investigation.

### Clinical and diagnostic relevance

The BPES phenotype is characterized by a typical eyelid malformation, including blepharophimosis, ptosis, epicanthus inversus and telecanthus, sometimes associated with POI. The phenotype can be distinguished from other syndromes that share one or more overlapping traits, such as Say-Barber-Biesecker variant of Ohdo syndrome (OMIM #603736) or Kaufman oculocerebrofacial syndrome (OMIM #244450), caused by variants in *KAT6B* and *UBE3B* respectively. Also, one case has been described with loss-of-function of NR2F2, resulting in a BPES-like phenotype (135).

However, to date *FOXL2* remains the only known disease gene underlying typical BPES. Importantly, penetrance for the eyelid phenotype in individuals carrying a heterozygous *FOXL2* variant is considered complete, as it is observed in all molecularly solved BPES patients. The only reported exception is a small *FOXL2* polyalanine expansion of +5 residues behaving as a recessive allele in a consanguineous Indian family (78). This variant likely represents the milder end of a length- and dosage-dependent pathogenic spectrum of *FOXL2* polyalanine expansions, as it was also found in a heterozygous state in a young, male BPES patient (104). In contrast, the penetrance of POI in BPES patients overall is considered reduced and age-related.

Molecular diagnosis remains necessary to genetically confirm BPES, particularly for clinical decision-making and accurate genetic counselling. Given the autosomal dominant inheritance pattern, genetic counselling is essential in BPES families, both for recurrence risk assessment and reproductive planning. This updated *FOXL2* variant database, including previously published and novel variants (**Tables S1, S2 & S3**), curated according to ACMG/AMP criteria, provides a key tool for clinical laboratories and clinicians to aid interpretation of variants identified in patients with BPES.

Because of the highly specific phenotype and complete penetrance of the eyelid malformation, we propose tailored rule specification of ACMG criterion PP4 for *FOXL2* variant classification in BPES (see also **supplementary text 1**):

- PP4: when BPES is clinically well described and assessed by a physician (distinguished from nonspecific ptosis).
- PP4 moderate: when the phenotype is explicitly confirmed by an expert ophthalmologist.
- PP4 strong: when, in addition, biological parameters such as FSH and AMH levels support a diagnosis of POI.

This phenotype-driven refinement will facilitate more accurate and consistent variant classification across diagnostic laboratories. In the supplementary text file (**supplementary text 1)**, we propose additional *FOXL2*-specific rule specifications for variant classification in BPES.

### Genotype-phenotype correlations

Previous studies have attempted to define a clear genotype-phenotype correlation in BPES. Truncating variants located upstream of the polyalanine tract tend to be linked to BPES type I (combining the eyelid malformation with POI), whereas polyalanine expansions appear more frequently associated with BPES type II (presenting only eyelid malformations) (118,136). Missense variants within the forkhead domain have generally been correlated with BPES type II (127,136). However, Li *et al.* (2021) suggested that missense variants situated in the central region of the forkhead domain may instead lead to the more severe BPES type I phenotype (26). Despite these observations, a consistent genotype-phenotype correlation across *FOXL2* variants has not yet been established, making the prediction of POI risk in young female patients with BPES challenging.

Also in this study, no clear genotype-phenotype correlations could be established, mainly due to the lack of appropriate clinical follow-up on fertility and endocrinological outcomes in female patients.

For many variants, only a single index patient has been identified, making it difficult to determine whether these variants are associated with an ovarian phenotype. This uncertainty often stems from the fact that some variants were found only in male patients, or in female patients who are still too young to evaluate ovarian (mal)function.

Notably, for several recurrent variants observed in multiple female BPES patients, including c.843_859dup [p.(Pro287Argfs*75)], c.655C>T [p.(Gln219*)], c.157C>T [p.(Gln53*)], c.650C>T [p.(Ser217Phe)] and c.672_701dup30 [p.(Ala225_Ala234dup)] (**Table S1**), the phenotype ranged from no ovarian involvement to primary or secondary amenorrhea, sometimes leading to infertility, demonstrating that this variability is probably not restricted to a specific variant type. In addition, all variant types and positions appeared capable of causing both BPES combined or not with POI, with varying ages of onset of POI.

We therefore propose that BPES should rather be considered a single condition displaying a continuum ranging from a fully penetrant eyelid phenotype to BPES with POI, which shows age-related penetrance in affected females. Accordingly, we emphasize that all female BPES patients, irrespective of their underlying variant, should receive appropriate endocrinological and fertility follow-up to enable timely detection and management of ovarian dysfunction.

### Diagnostic perspective for patients with BPES

Currently, the diagnostic strategy for BPES relies on targeted *FOXL2* sequencing in patients with a typical eyelid phenotype, complemented by CNV analysis of the coding region and upstream non-coding regulatory region. This approach allows the majority of affected individuals to be molecularly confirmed. Nevertheless, approximately 12% of clinically diagnosed BPES patients remain without a molecular diagnosis (14). One striking example in our cohort was a multigenerational family of Polynesian descent that remained unsolved for many years. As no female family members had offspring, BPES with POI was suspected. Standard diagnostic testing with targeted Sanger sequencing of the *FOXL2* coding region and MLPA failed to identify a pathogenic variant. Whole-genome sequencing (WGS) of multiple family members eventually revealed the most recurrent polyalanine expansion, c.672_701dup (p.[Ala225_Ala234dup]) to co-segregate with disease. The earlier negative results were likely explained by allelic drop-out in tested individuals. As this expansion represents a major fraction of *FOXL2* variants, the case illustrates the limitations of current first-tier approaches and highlights the need for more specifically targeted methodologies. Bunyan *et al*. (2019) described a fluorescent PCR-based assay for detecting *FOXL2* polyalanine expansions, providing a reliable and cost-effective solution for identifying polyalanine expansions (31).

Beyond repeat expansions, additional classes of pathogenic variation may contribute to the unresolved fraction of BPES cases. Short-read and long-read WGS, as well as optical genome mapping, offer the ability to detect cryptic and complex SVs, including balanced rearrangements and long-range regulatory disruptions affecting the *FOXL2 cis*-regulatory landscape, which are not captured by current targeted assays. Moreover, most diagnostic workflows do not specifically interrogate the *FOXL2* promoter, raising the possibility that pathogenic deletions or regulatory alterations in this region remain undetected and may also account for part of the missing heritability.

Together, the integration of repeat-sensitive assays, long-read sequencing, and genome-wide SV detection approaches has the potential to substantially improve diagnostic yield, reduce the proportion of unsolved BPES cases, and further refine the understanding of *FOXL2 cis*-regulatory mechanisms, ultimately improving patient care and genetic counseling.

## 5. Conclusion

In this study, we present a comprehensive overview and ACMG/AMP-based classification of virtually all reported *FOXL2* variants associated with BPES up to December 2024 and expand the variant spectrum by reporting 76 novel genetic defects. We showed that genotype-phenotype correlations for POI are not straightforward, making prediction of POI risk in young female patients with BPES particularly challenging. This is partly due to the lack of appropriate longitudinal clinical follow-up, which is particularly relevant for prepubertal female patients. Here, we also propose that BPES should not be subclassified in two distinct subtypes, but rather as a condition with a fully penetrant eyelid malformation, consistently present in all patients, with reduced penetrance and variable expressivity of POI in affected females. Looking forward, improved diagnostic strategies, including (targeted) long-read sequencing and optical genome mapping to resolve variants in the polyalanine tract and short-read and long-read WGS to capture complex SVs and non-coding alterations, will be essential to increase the diagnostic yield and ultimately improve precision medicine in BPES.

## Data Availability

All data produced in the present work are contained in the manuscript.

## 7. Author contributions

Charlotte Matton: conceptualization, data curation, formal analysis, investigation, visualization, writing of the original draft. Julie Van De Velde: data curation, formal analysis, investigation, review and editing. Marieke De Bruyne: data curation, formal analysis, review and editing. Stijn Van De Sompele: data curation, formal analysis, review and editing. Sally Hooghe: data curation, formal analysis, review and editing. Miriam Bauwens: review and editing. Hannes Syryn: review and editing. Eva D’haene: review and editing. Annelies Dheedene: formal analysis, review and editing. Martine Cools: review and editing. Wendy Watkins: formal analysis, review and editing. Shoko Komatsuzaki: formal analysis, resources, review and editing. Ewelina Preizner-Rzucidło: formal analysis, resources, review and editing. Alison Ross: formal analysis, resources, review and editing. Christine Armstrong: data curation, formal analysis, review and editing. David Bunyan: formal analysis, resources, review and editing. Andrew Shelling: formal analysis, resources, review and editing. Andrea Vincent: formal analysis, resources, review and editing. Sasha Vermeer: formal analysis, resources, review and editing. Hannah Verdin: data curation, formal analysis, review and editing. Elfride De Baere: conceptualization, funding acquisition, supervision, review and editing.

## 9. Acknowledgements

The authors would like to thank Elien Wallican and Arend Segaert for their contribution in context of their bachelor dissertations. ChatGPT was used as language model to assist with text editing. E.D.B. is member of ERN-EYE and E.D.B. and M.C. of EndoERN (Framework Partnership Agreement No 739534-ERN-EYE and EndoERN).

## 11. Supplementary files

**Table S1** All identified frameshift, nonsense, missense, in-frame indel, and stop-loss variants within the *FOXL2* coding region that are associated with a blepharophimosis, ptosis, epicanthus inversus syndrome (BPES) phenotype.

For each variant, the table provides the genomic coordinates, predicted protein effect, and variant classification according to the ACMG/AMP framework. The applied evidence criteria and corresponding arguments supporting each classification are also included.

**Table S2** Overview of all structural variants (deletions) identified in the *FOXL2* locus and its flanking regions, including intragenic *FOXL2* deletions, combined *FOXL2*-*ATR* deletions, and deletions located up- or downstream of *FOXL2*, including genomic coordinates (when available) and the associated phenotype.

**Table S3** Overview of all translocations that disrupt or reposition the *FOXL2* locus and are associated with a blepharophimosis, ptosis, epicanthus inversus syndrome (BPES) phenotype. These data highlight the contribution of structural chromosomal rearrangements to BPES and illustrate how disruption of *FOXL2* or its regulatory landscape can lead to variable or more severe clinical manifestations.

**Table S4** Overview of the primers that are used for Sanger sequencing in BPES, covering the entire *FOXL2* gene.

**Supplementary text 1** explains how genetic variants were classified using an ACMG/AMP-based framework that considers seven types of evidence (population, genotype/phenotype, databases, computational, functional, segregation, and allelic data) to determine pathogenicity.

## References

1. Cocquet J, De Baere E, Gareil M, Pannetier M, Xia X, Fellous M, et al. Structure, evolution and expression of the FOXL2 transcription unit. Cytogenet Genome Res [Internet]. 2003 [cited 2022 Nov 24];101(3–4):206–11. Available from: https://www.karger.com/Article/FullText/74338

2. Crisponi L, Deiana M, Loi A, Chiappe F, Uda M, Amati P, et al. The putative forkhead transcription factor FOXL2 is mutated in blepharophimosis/ptosis/epicanthus inversus syndrome. Nature Genetics 2001 27:2 [Internet]. 2001 [cited 2022 Oct 5];27(2):159–66. Available from: https://www.nature.com/articles/ng0201_159

3. De Baere E, Dixon MJ, Small KW, Jabs EW, Leroy BP, Devriendt K, et al. Spectrum of FOXL2 gene mutations in blepharophimosis-ptosis-epicanthus inversus (BPES) families demonstrates a genotype–phenotype correlation. Hum Mol Genet [Internet]. 2001 Jul 15 [cited 2022 Oct 14];10(15):1591–600. Available from: https://academic.oup.com/hmg/article/10/15/1591/587684

4. Lehmann OJ, Sowden JC, Carlsson P, Jordan T, Bhattacharya SS. Fox’s in development and disease. Vol. 19, Trends in Genetics. Elsevier Ltd; 2003. p. 339–44.

5. Zhang Y, Kao WWY, Pelosi E, Schlessinger D, Liu CY. Notch gain of function in mouse periocular mesenchyme downregulates FoxL2 and impairs eyelid levator muscle formation, leading to congenital blepharophimosis. J Cell Sci [Internet]. 2011 Aug 8 [cited 2022 Nov 29];124(15):2561. Available from: /pmc/articles/PMC3138700/

6. Tucker EJ. The Genetics and Biology of FOXL2. Sexual Development [Internet]. 2022 [cited 2022 Oct 17];16(2–3):184–93. Available from: https://www.karger.com/Article/FullText/519836

7. Schmidt D, Ovitt CE, Anlag K, Fehsenfeld S, Gredsted L, Treier AC, et al. The murine winged-helix transcription factor Foxl2 is required for granulosa cell differentiation and ovary maintenance. Development [Internet]. 2004 Feb 15 [cited 2022 Oct 5];131(4):933–42. Available from: https://journals.biologists.com/dev/article/131/4/933/42626/The-murine-winged-helix-transcription-factor-Foxl2

8. Uda M, Ottolenghi C, Crisponi L, Garcia JE, Deiana M, Kimber W, et al. Foxl2 disruption causes mouse ovarian failure by pervasive blockage of follicle development. Hum Mol Genet [Internet]. 2004 Jun 1 [cited 2022 Oct 5];13(11):1171–81. Available from: https://academic.oup.com/hmg/article/13/11/1171/699061

9. Ellsworth BS, Egashira N, Haller JL, Butts DL, Cocquet J, Clay CM, et al. FOXL2 in the Pituitary: Molecular, Genetic, and Developmental Analysis. Molecular Endocrinology [Internet]. 2006 Nov 1 [cited 2022 Nov 24];20(11):2796–805. Available from: https://academic.oup.com/mend/article/20/11/2796/2738404

10. Castets S, Roucher-Boulez F, Saveanu A, Mallet-Motak D, Chabre O, Amati-Bonneau P, et al. Hypopituitarism in Patients with Blepharophimosis and FOXL2 Mutations. Horm Res Paediatr. 2020 Jul 1;93(1):30–9.

11. Governini L, Carrarelli P, Rocha ALL, Leo V De, Luddi A, Arcuri F, et al. FOXL2 in human endometrium: Hyperexpressed in endometriosis. Reproductive Sciences. 2014 Oct 11;21(10):1249–55.

12. Zhang B, Li SJ, Yuan H, Cong SS, Zhao SJ, Yang XJ. FOXL2 Knockdown Inhibits the Progression of Endometriosis. American Journal of Reproductive Immunology. 2025 Jan 1;93(1).

13. Uhlenhaut NH, Jakob S, Anlag K, Eisenberger T, Sekido R, Kress J, et al. Somatic Sex Reprogramming of Adult Ovaries to Testes by FOXL2 Ablation. Cell. 2009 Dec 11;139(6):1130–42.

14. Verdin H, Matton C, Baere E De. Blepharophimosis, Ptosis, and Epicanthus Inversus Syndrome. 2022 Mar 10 [cited 2022 Oct 17];1–19. Available from: https://www.ncbi.nlm.nih.gov/books/NBK1441/

15. Owens N, Hadley RC, Kloepfer HW. Hereditary blepharophimosis, ptosis, and epicanthus inversus. J Int Coll Surg [Internet]. 1960 May [cited 2022 Oct 5];33:558–74. Available from: https://pubmed.ncbi.nlm.nih.gov/14429566/

16. Shah SP, Köbel M, Senz J, Morin RD, Clarke BA, Wiegand KC, et al. Mutation of FOXL2 in Granulosa-Cell Tumors of the Ovary. New England Journal of Medicine [Internet]. 2009 Jun 25 [cited 2022 Nov 24];360(26):2719–29. Available from: https://www.nejm.org/doi/10.1056/NEJMoa0902542

17. Beysen D, De Paepe A, De Baere E. Human Mutation MUTATION UPDATE FOXL2 Mutations and Genomic Rearrangements in BPES. Hum Mutat [Internet]. 2009 [cited 2023 Jan 2];30:158–69. Available from: https://onlinelibrary.wiley.com/doi/10.1002/humu.20807

18. D’Haene B, Nevado J, Pugeat M, Pierquin G, Lowry RB, Reardon W, et al. FOXL2 copy number changes in the molecular pathogenesis of BPES: unique cohort of 17 deletions. Hum Mutat [Internet]. 2010 May 1 [cited 2022 Oct 10];31(5):E1332–47. Available from: https://onlinelibrary.wiley.com/doi/full/10.1002/humu.21233

19. Beysen D, Raes J, Leroy BP, Lucassen A, Yates JRW, Clayton-Smith J, et al. Deletions Involving Long-Range Conserved Nongenic Sequences Upstream and Downstream of FOXL2 as a Novel Disease-Causing Mechanism in Blepharophimosis Syndrome. Am J Hum Genet. 2005;77:205–18.

20. Verdin H, D’haene B, Beysen D, Novikova Y, Menten B, Sante T, et al. Microhomology-Mediated Mechanisms Underlie Non-Recurrent Disease-Causing Microdeletions of the FOXL2 Gene or Its Regulatory Domain. PLoS Genet [Internet]. 2013 [cited 2022 Oct 15];9(3). Available from: /pmc/articles/PMC3597517/

21. Richards Chair S, Bick ACMG D, Das ACMG S, Gastier-Foster AMP J, Bick D, Das S, et al. Standards and Guidelines for the Interpretation of Sequence Variants: A Joint Consensus Recommendation of the American College of Medical Genetics and Genomics and the Association for Molecular Pathology. Genetics in Medicine [Internet]. 2015 [cited 2024 May 2];17(5):405–24. Available from: http://www.nature.com/authors/editorial_policies/license.html#terms

22. Tavtigian S V., Greenblatt MS, Harrison SM, Nussbaum RL, Prabhu SA, Boucher KM, et al. Modeling the ACMG/AMP variant classification guidelines as a Bayesian classification framework. Genetics in Medicine [Internet]. 2018 Sep 1 [cited 2025 Oct 14];20(9):1054–60. Available from: https://pubmed.ncbi.nlm.nih.gov/29300386/

23. Hongbo W, Zhang Z. A Case Report of Primary Infertility With BPES Syndrome With FOXL2 Gene Mutation and PADI6 Gene Mutation. Journal of Clinical Medicine & Surgery; Case Report [Internet]. 2022 [cited 2022 Oct 14];1(1):1–9. Available from: https://jcmsr.org//

24. Meng T, Zhang W, Zhang R, Li J, Gao Y, Qin Y, et al. Ovarian Reserve and ART Outcomes in Blepharophimosis-Ptosis-Epicanthus Inversus Syndrome Patients With FOXL2 Mutations. Front Endocrinol (Lausanne) [Internet]. 2022 Apr 28 [cited 2022 Sep 21];13:1. Available from: https://www.frontiersin.org/articles/10.3389/fendo.2022.829153/full

25. Eskenazi S, Bachelot A, Hugon-Rodin J, Plu-Bureau G, Gompel A, Catteau-Jonard S, et al. Next Generation Sequencing Should Be Proposed to Every Woman With “Idiopathic” Primary Ovarian Insufficiency. J Endocr Soc [Internet]. 2021;5(7):1–10. Available from: https://academic.oup.com/jes1

26. Li F, Chen H, Wang Y, Yang J, Zhou Y, Song X, et al. Functional Studies of Novel FOXL2 Variants in Chinese Families With Blepharophimosis–Ptosis–Epicanthus Inversus Syndrome. Front Genet. 2021 Mar 16;12:224.

27. Rong WN, Yang W, Yuan SQ, Sheng XL. Identification of a novel FOXL2 mutation in a fourth-generation Chinese family with blepharophimosis-ptosis-epicanthus inversus syndrome. Int J Ophthalmol [Internet]. 2021 Apr 18;14(4):504–9. Available from: http://ies.ijo.cn/gjyken/ch/reader/view_abstract.aspx?file_no=20210404&flag=1

28. Zheng B, Seltzsam S, Wang C, Schierbaum L, Schneider S, Wu CHW, et al. Whole-exome sequencing identifies FOXL2, FOXA2 and FOXA3 as candidate genes for monogenic congenital anomalies of the kidneys and urinary tract. Nephrology Dialysis Transplantation [Internet]. 2022 Sep 22 [cited 2022 Oct 14];37(10):1833–43. Available from: https://academic.oup.com/ndt/article/37/10/1833/6362937

29. Wang S, Ge S, Zhuang A. A Novel Forkhead Box L2 Missense Mutation, c.1068G>C, in a Chinese Family with Blepharophimosis/Ptosis/ Epicanthus Inversus Syndrome. Journal of Craniofacial Surgery [Internet]. 2022 May 1 [cited 2022 Oct 14];33(3):E238–40. Available from: https://journals.lww.com/jcraniofacialsurgery/Fulltext/2022/05000/A_Novel_Forkhead_Box_L2_Missense_Mutation,.71.aspx

30. Hu J, Ke H, Luo W, Yang Y, Liu H, Li G, et al. A novel FOXL2 mutation in two infertile patients with blepharophimosis–ptosis–epicanthus inversus syndrome. J Assist Reprod Genet [Internet]. 2020 Jan 1 [cited 2022 Oct 14];37(1):223–9. Available from: https://link.springer.com/article/10.1007/s10815-019-01651-2

31. Bunyan DJ, Thomas NS. Screening of a large cohort of blepharophimosis, ptosis, and epicanthus inversus syndrome patients reveals a very strong paternal inheritance bias and a wide spectrum of novel FOXL2 mutations. Eur J Med Genet. 2019 Jul 1;62(7).

32. Bertini V, Valetto A, Baldinotti F, Azzarà A, Cambi F, Toschi B, et al. Blepharophimosis, Ptosis, Epicanthus Inversus Syndrome: New Report with a 197-kb Deletion Upstream of FOXL2 and Review of the Literature. Mol Syndromol [Internet]. 2019 May 1 [cited 2022 Oct 14];10(3):147–53. Available from: https://www.karger.com/Article/FullText/497092

33. Chacón-Camacho OF, Salgado-Medina A, Alcaraz-Lares N, López-Moreno D, Barragán-Arévalo T, Nava-Castañeda A, et al. Clinical characterization and identification of five novel FOXL2 pathogenic variants in a cohort of 12 Mexican subjects with the syndrome of blepharophimosis-ptosis-epicanthus inversus. Gene. 2019 Jul 20;706:62–8.

34. Grzechocińska B, Warzecha D, Wypchło M, Ploski R, Wielgoś M. Premature ovarian insufficiency as a variable feature of blepharophimosis, ptosis, and epicanthus inversus syndrome associated with c.223C > T p.(Leu75Phe) FOXL2 mutation: a case report. BMC Med Genet [Internet]. 2019 Jul 31 [cited 2022 Oct 5];20(1). Available from: /pmc/articles/PMC6670140/

35. Bouman A, Van Haelst M, Van Spaendonk R. Blepharophimosis-ptosis-epicanthus inversus syndrome caused by a 54-kb microdeletion in a FOXL2 cis-regulatory element. Clin Dysmorphol [Internet]. 2018 [cited 2022 Oct 14];27(2):58–62. Available from: https://journals.lww.com/clindysmorphol/Fulltext/2018/04000/Blepharophimosis_ptosis_epicanthus_inversus.8.aspx

36. Yang XW, He WB, Gong F, Li W, Li XR, Zhong CG, et al. Novel FOXL2 mutations cause blepharophimosis-ptosis-epicanthus inversus syndrome with premature ovarian insufficiency. 2018 [cited 2022 Sep 29]; Available from: www.mutationtaster.org/

37. Zhou L, Wang J, Wang T. Functional study on new FOXL2 mutations found in Chinese patients with blepharophimosis, ptosis, epicanthus inversus syndrome. BMC Med Genet [Internet]. 2018 Jul 20 [cited 2022 Oct 14];19(1):1–7. Available from: https://bmcmedgenet.biomedcentral.com/articles/10.1186/s12881-018-0631-8

38. Cheng H, Wang T, Wang G, Wang J, Shen L, Han M, et al. Analysis of FOXL2 gene mutation and genotype-phenotype correlation in a Chinese pedigree affected with blepharophimosis-ptosis-epicanthus inversus syndrome. Zhonghua Yi Xue Yi Chuan Xue Za Zhi [Internet]. 2018 Aug 1 [cited 2022 Oct 14];35(4):515–7. Available from: https://pubmed.ncbi.nlm.nih.gov/30098246/

39. Li F, Chai P, Fan J, Wang X, Lu W, Li J, et al. A Novel FOXL2 Mutation Implying Blepharophimosis-Ptosis-Epicanthus Inversus Syndrome Type I. Cellular Physiology and Biochemistry [Internet]. 2018 Feb 1 [cited 2022 Sep 29];45(1):203–11. Available from: https://www.karger.com/Article/FullText/486358

40. Li H, Gu Y. Genetic and functional analyses of two missense mutations in the transcription factor FOXL2 in two Chinese families with blepharophimosis-ptosis-epicanthus inversus syndrome. Genet Test Mol Biomarkers [Internet]. 2018 Oct 1 [cited 2022 Oct 14];22(10):585–92. Available from: https://www.liebertpub.com/doi/10.1089/gtmb.2018.0064

41. Niu BB, Tang N, Xu Q, Chai PW. Genomic Disruption of FOXL2 in Blepharophimosis-Ptosis-Epicanthus Inversus Syndrome Type 2: A Novel Deletion-Insertion Compound Mutation. Chin Med J (Engl) [Internet]. 2018 Oct 10 [cited 2022 Oct 14];131(19):2380. Available from: /pmc/articles/PMC6166469/

42. Yang L, Li T, Xing Y. Identification of a novel FOXL2 Mutation in a single family with both types of Blepharophimosis-ptosis-epicanthus inversus syndrome. Mol Med Rep [Internet]. 2017 Oct 1 [cited 2022 Oct 14];16(4):5529–32. Available from: http://www.spandidos-publications.com/10.3892/mmr.2017.7226/abstract

43. Duarte AF, Akaishi PMS, de Molfetta GA, Chodraui-Filho S, Cintra M, Toscano A, et al. Lacrimal Gland Involvement in Blepharophimosis-Ptosis-Epicanthus Inversus Syndrome. Ophthalmology. 2017 Mar 1;124(3):399–406.

44. Yang X, Li W, Du J, Yuan S, He W, Zhang Q, et al. Analysis of FOXL2 gene mutations in 5 families affected with blepharophimosis, ptosis and epicanthus inversus syndrome. Zhonghua Yi Xue Yi Chuan Xue Za Zhi [Internet]. 2017 Jun 1 [cited 2022 Sep 26];34(3):342–6. Available from: https://pubmed.ncbi.nlm.nih.gov/28604951/

45. Chai P, Li F, Fan J, Jia R, Fan X. Functional Analysis of a Novel FOXL2 Indel Mutation in Chinese Families with Blepharophimosis-Ptosis-Epicanthus Inversus Syndrome Type I. Int J Biol Sci [Internet]. 2017;13. Available from: http://www.ijbs.com

46. Krepelova A, Simandlova M, Vlckova M, Kuthan P, Vincent AL, Liskova P. Analysis of FOXL2 detects three novel mutations and an atypical phenotype of blepharophimosis-ptosis-epicanthus inversus syndrome. 2016; Available from: www.hgvs.org/mutnomen/

47. Tan H, Yang P, Li H, Pan Q, Liang D, Wu L. A novel FOXL2 mutation in a Chinese family with blepharophimosis, ptosis, epicanthus inversus syndrome. Nature Publishing Group [Internet]. 2015;2. Available from: www.nature.com/hgv

48. Xue M, Zheng J, Zhou Q, Fielding Hejtmancik J, Wang Y, Li S. Novel FOXL2 mutations in two Chinese families with blepharophimosis-ptosis-epicanthus inversus syndrome. 2015; Available from: http://provean.jcvi.org/index.php

49. Settas N, Anapliotou M, Kanavakis E, Fryssira H, Sofocleous C, Dacou-Voutetakis C, et al. A novel FOXL2 gene mutation and BMP15 variants in a woman with primary ovarian insufficiency and blepharophimosis-ptosis-epicanthus inversus syndrome. Menopause [Internet]. 2015 Oct 27 [cited 2022 Sep 28];22(11):1264–8. Available from: https://journals.lww.com/menopausejournal/Fulltext/2015/11000/A_novel_FOXL2_gene_mutation_and_BMP15_variants_in.16.aspx

50. Nuovo S, Passeri · M, Di Benedetto · E, Calanchini · M, Meldolesi · I, Di Giacomo · M C, et al. Characterization of endocrine features and genotype-phenotypes correlations in blepharophimosis-ptosis-epicanthus inversus syndrome type 1. J Endocrinol Invest [Internet]. 2016 [cited 2022 Sep 23];39:227–33. Available from: http://medgen.ugent.be/

51. Martinez-Aguayo A, Poggi H, Cattani A, Molina M, Romeo E, Lagos M. A novel insertion in the FOXL2 gene in a Chilean patient with blepharophimosis ptosis epicanthus inversus syndrome type i. Journal of Pediatric Endocrinology and Metabolism [Internet]. 2014 Jan 1 [cited 2022 Oct 14];27(1–2):181–4. Available from: https://www.degruyter.com/document/doi/10.1515/jpem-2013-0219/html

52. Gulati R, Verdin H, Halanaik D, Bhat BV, De Baere E. Co-occurrence of congenital hydronephrosis and FOXL2-associated blepharophimosis, ptosis, epicanthus inversus syndrome (BPES). Eur J Med Genet. 2014 Oct 1;57(10):576–8.

53. Jiang H, Huang X, Su Z, Rao L, Wu S, Zhang T, et al. Genetic analysis of the forkhead transcriptional factor 2 gene in three Chinese families with blepharophimosis syndrome. Mol Vis [Internet]. 2013 Feb 20 [cited 2022 Oct 14];19:418. Available from: /pmc/articles/PMC3580973/

54. Zhang L, Wang L, Han R, Guan L, Fan B, Liu M, et al. Identification of the forkhead transcriptional factor 2 (FOXL2) gene mutations in four Chinese families with blepharophimosis syndrome. Mol Vis [Internet]. 2013 Nov 16 [cited 2022 Oct 14];19:2298. Available from: /pmc/articles/PMC3834601/

55. Fan J, Zhou Y, Huang X, Zhang L, Yao Y, Song X, et al. The combination of polyalanine expansion mutation and a novel missense substitution in transcription factor FOXL2 leads to different ovarian phenotypes in blepharophimosis–ptosis–epicanthus inversus syndrome (BPES) patients. Human Reproduction [Internet]. 2012 Nov 1 [cited 2022 Oct 14];27(11):3347–57. Available from: https://academic.oup.com/humrep/article/27/11/3347/812262

56. Haghighi A, Verdin H, Haghighi-Kakhki H, Piri N, Gohari NS, De Baere E. Missense mutation outside the forkhead domain of FOXL2 causes a severe form of BPES type II. 2012 [cited 2022 Sep 23]; Available from: http://www.molvis.org/molvis/v18/a24

57. Zahanova S, Meaney B, Łabieniec B, Verdin H, De Baere E, Nowaczyk MJM. Blepharophimosis-ptosis-epicanthus inversus syndrome plus: Deletion 3q22.3q23 in a patient with characteristic facial features and with genital anomalies, spastic diplegia, and speech delay. Clin Dysmorphol. 2012 Jan;21(1):48–52.

58. González-González C, García-Hoyos M, Hernaez Calzón R, Arroyo Díaz C, González Fanego C, Lorda Sánchez I, et al. Microdeletion found by array-CGH in girl with Blepharophimosis syndrome and apparently balanced translocation t(3;15)(q23;q25). 10.3109/138168102011634879 [Internet]. 2012 [cited 2022 Sep 28];33(2):107–10. Available from: https://www.tandfonline.com/doi/abs/10.3109/13816810.2011.634879

59. Hu S, Guo J, Wang B, Wang J, Zhou Z, Zhou G, et al. Genetic analysis of the FOXL2 gene using quantitative real-time PCR in Chinese patients with blepharophimosis-ptosis-epicanthus inversus syndrome. Mol Vis [Internet]. 2011 [cited 2022 Sep 28];17:436. Available from: /pmc/articles/PMC3038209/

60. Kaur I, Hussain A, Naik MN, Murthy R, Honavar SG. Mutation spectrum of Fork-Head Transcriptional Factor Gene (FOXL2) in Indian Blepharophimosis Ptosis Epicanthus Inversus Syndrome (BPES) patients. British Journal of Ophthalmology [Internet]. 2011 Jun 1 [cited 2022 Sep 28];95(6):881–6. Available from: https://bjo.bmj.com/content/95/6/881

61. Tang S, Wang X, Lin L, Sun Y, Wang Y, Yu H. Mutation analysis of the FOXL2 gene in Chinese patients with blepharophimosis-ptosis-epicanthus inversus syndrome. Mutagenesis. 2006 Jan;21(1):35–9.

62. Fan JY, Han B, Qiao J, Liu BL, Ji YR, Ge SF, et al. Functional study on a novel missense mutation of the transcription factor FOXL2 causes blepharophimosis-ptosis-epicanthus inversus syndrome (BPES). Mutagenesis [Internet]. 2011 [cited 2022 Sep 28];26(2):283–9. Available from: https://academic.oup.com/mutage/article/26/2/283/1091378

63. Chouchene I, Derouiche K, Chaabouni A, Cherif L, Amouri A, Largueche L, et al. Identification of a novel mutation in FOXL2 gene that leads to blepharophimosis ptosis epicanthus inversus and telecanthus syndrome in a Tunisian consanguineous family. Genet Test Mol Biomarkers [Internet]. 2010 Feb 1 [cited 2022 Sep 27];14(1):145–8. Available from: https://www.liebertpub.com/doi/10.1089/gtmb.2009.0091

64. Kraoua L, Chaabouni M, Trabelsi M, Chelly I, Maazoul F, Ben Abdallah N, et al. FOXL2 mutations in Tunisian patients with blepharophimosis-ptosis-epicanthus inversus syndrome. Clin Genet. 2010 Jun;77(6):601–3.

65. Lin W De, Chou IC, Lee NC, Wang CH, Hwu WL, Lin SP, et al. FOXL2 mutations in Taiwanese patients with blepharophimosis, ptosis, epicanthus inversus syndrome. Clin Chem Lab Med [Internet]. 2010 Apr 1 [cited 2022 Oct 14];48(4):485–8. Available from: https://www.degruyter.com/document/doi/10.1515/CCLM.2010.100/html

66. Zhou Z min, Liang D sheng, Quan Y, Xue J jie, Li H yan, Xia X bo, et al. Deletion and mutation analysis to FOXL2 in blepharophimosis-ptosis-epicanthus inversus syndrome. Zhonghua Yan Ke Za Zhi [Internet]. 2010 Jun;46(6):532–6. Available from: http://www.ncbi.nlm.nih.gov/pubmed/21055199

67. Ni F, Wen Q, Wang B, Zhou S, Wang J, Mu Y, et al. Mutation analysis of FOXL2 gene in Chinese patients with premature ovarian failure. Gynecological Endocrinology. 2010 Apr;26(4):246–9.

68. D’haene B, Attanasio C, Beysen D, Dostie J, Lemire E, Bouchard P, et al. Disease-Causing 7.4 kb Cis-Regulatory Deletion Disrupting Conserved Non-Coding Sequences and Their Interaction with the FOXL2 Promotor: Implications for Mutation Screening. Horwitz MS, editor. PLoS Genet [Internet]. 2009 Jun 19 [cited 2022 Sep 27];5(6):e1000522. Available from: https://dx.plos.org/10.1371/journal.pgen.1000522

69. Méduri G, Bachelot A, Duflos C, Bständig B, Poirot C, Genestie C, et al. FOXL2 mutations lead to different ovarian phenotypes in BPES patients: Case Report. Human Reproduction [Internet]. 2010 Jan 1 [cited 2022 Oct 14];25(1):235–43. Available from: https://academic.oup.com/humrep/article/25/1/235/696004

70. Li D, Zeng W, Tao J, Li S, Liang C, Chen X, et al. Mutations of the transcription factor FOXL2 gene in Chinese patients with blepharophimosis-ptosis-epicanthus inversus syndrome. Genet Test Mol Biomarkers. 2009;13(2):257–68.

71. Xu Y, Lei H, Dong H, Zhang L, Qin Q, Gao J, et al. FOXL2 gene mutations and blepharophimosis–ptosis–epicanthus inversus syndrome (BPES): a novel mutation detected in a Chinese family and a statistic model for summarizing previous reported records. Mutagenesis [Internet]. 2009 Sep 1 [cited 2022 Oct 14];24(5):447–53. Available from: https://academic.oup.com/mutage/article/24/5/447/1747500

72. Corrêa FJS, Tavares AB, Pereira RW, Abrão MS. A new FOXL2 gene mutation in a woman with premature ovarian failure and sporadic blepharophimosis-ptosis-epicanthus inversus syndrome. Fertil Steril. 2010 Feb 1;93(3):1006.e3-1006.e6.

73. Beysen D, De Jaegere S, Amor D, Bouchard P, Christin-Maitre S, Fellous M, et al. Identification of 34 novel and 56 known FOXL2 mutations in patients with blepharophimosis syndrome. Hum Mutat. 2008 Nov;29(11).

74. Nallathambi J, Laissue P, Batista F, Benayoun BA, Lesaffre C, Moumné L, et al. Differential functional effects of novel mutations of the transcription factor FOXL2 in BPES patients. Hum Mutat. 2008;29(8):E123–31.

75. Tzschach A, Kelbova C, Weidensee S, Peters H, Ropers HH, Ullmann R, et al. Blepharophimosis-Ptosis-Epicanthus Inversus Syndrome in a Girl with Chromosome Translocation t(2;3)(q33;q23). 10.1080/13816810701867615 [Internet]. 2009 Mar [cited 2022 Oct 14];29(1):37–40. Available from: https://www.tandfonline.com/doi/abs/10.1080/13816810701867615

76. Leon-Mateos A, Ginarte M, Ruiz-Ponte C, Carracedo A, Toribio J. Blepharophimosis?ptosis?epicanthus inversus syndrome (BPES). Int J Dermatol [Internet]. 2007 Jan;46(1):61–3. Available from: https://onlinelibrary.wiley.com/doi/10.1111/j.1365-4632.2007.03066.x

77. Wang J, Liu J, Zhang Q. FOXL2 mutations in Chinese patients with blepharophimosis-ptosis-epicanthus inversus syndrome. Mol Vis [Internet]. 2007 Jan 26 [cited 2022 Sep 23];13:108–13. Available from: http://www.ncbi.nlm.nih.gov/pubmed/17277738

78. Nallathambi J, Neethirajan G, Usha K, Jitendra J, De Baere E, Sundaresan P. FOXL2 mutations in Indian families with blepharophimosis-ptosis-epicanthus inversus syndrome. J Genet. 2007 Aug;86(2):165–8.

79. Or SFJ, Tong MFT, Lo FMI, Lam TSS. Three novel FOXL2 gene mutations in Chinese patients with blepharophimosis-ptosis-epicanthus inversus syndrome. Chin Med J (Engl). 2006 Jan 5;119(1):49–52.

80. Nallathambi J, Moumné L, De Baere E, Beysen D, Usha K, Sundaresan P, et al. A novel polyalanine expansion in FOXL2: the Wrst evidence for a recessive form of the blepharophimosis syndrome (BPES) associated with ovarian dysfunction. Hum Genet [Internet]. 2007 [cited 2022 Sep 23];121:107–12. Available from: http://www.medgen.

81. Mari F, Giachino D, Russo L, Pilia G, Ariani F, Scala E, et al. Blepharophimosis, Ptosis, and Epicanthus Inversus Syndrome: Clinical and Molecular Analysis of a Case. Journal of American Association for Pediatric Ophthalmology and Strabismus. 2006 Jun 1;10(3):279–80.

82. De Ru MH, Gille JJP, Nieuwint AWM, Bijlsma JB, Van Der Blij JF, Van Hagen JM. Interstitial deletion in 3q in a patient with blepharophimosis-ptosis-epicanthus inversus syndrome (BPES) and microcephaly, mild mental retardation and growth delay: clinical report and review of the literature. Am J Med Genet A [Internet]. 2005 Aug 15 [cited 2022 Sep 22];137(1):81–7. Available from: https://pubmed.ncbi.nlm.nih.gov/16015581/

83. Raile K, Stobbe H, Tröbs RB, Kiess W, Pfäffle R. A new heterozygous mutation of the FOXL2 gene is associated with a large ovarian cyst and ovarian dysfunction in an adolescent girl with blepharophimosis/ptosis/epicanthus inversus syndrome. Eur J Endocrinol. 2005 Sep;153(3):353–8.

84. Kumar A, Babu M, Raghunath A, Venkatesh CP. Genetic analysis of a five generation Indian family with BPES: a novel missense mutation (p.Y215C). Mol Vis. 2004 Jul 9;10:445–9.

85. de Baere E, Beysen D, Oley C, Lorenz B, Cocquet J, de Sutter P, et al. FOXL2 and BPES: Mutational Hotspots, Phenotypic Variability, and Revision of the Genotype-Phenotype Correlation. Am J Hum Genet. 2003;72:478–87.

86. Udar N, Yellore V, Chalukya M, Yelchits S, Silva-Garcia R, Anderson R, et al. Comparative analysis of the FOXL2 gene and characterization of mutations in BPES patients. Hum Mutat. 2003;22(3):222–8.

87. Cha SC, Jang YS, Lee JH, Kim HK, Kim SC, Kim S, et al. Mutational analysis of forkhead transcriptional factor 2 (FOXL2) in Korean patients with blepharophimosis-ptosis-epicanthus inversus syndrome. Clin Genet [Internet]. 2003 Dec 18 [cited 2022 Sep 20];64(6):485–90. Available from: http://www.ncbi.nlm.nih.gov/pubmed/14986827

88. Fokstuen S, Antonarakis SE, Blouin JL. FOXL2-mutations in blepharophimosis-ptosis - Epicanthus inversus syndrome (BPES); challenges for genetic counseling in female patients. Am J Med Genet. 2003 Mar 1;117 A(2):143–6.

89. Dollfus H, Stoetzel C, Riehm S, Lahlou Boukoffa W, Bediard Boulaned F, Quillet R, et al. Sporadic and familial blepharophimosis-ptosis-epicanthus inversus syndrome: FOXL2 mutation screen and MRI study of the superior levator eyelid muscle. Clin Genet. 2003 Feb 1;63(2):117–20.

90. Harris SE, Chand AL, Winship IM, Gersak K, Aittomäki K, Shelling AN. Identification of novel mutations in FOXL2 associated with premature ovarian failure. Mol Hum Reprod [Internet]. 2002 Aug 1 [cited 2022 Oct 14];8(8):729–33. Available from: https://academic.oup.com/molehr/article/8/8/729/992187

91. Kosaki K, Ogata T, Kosaki R, Sato S, Matsuo N. A novel mutation in the FOXL2 gene in a patient with blepharophimosis syndrome: Differential role of the polyalanine tract in the development of the ovary and the eyelid. 10.1076/opge231432202 [Internet]. 2009 [cited 2022 Oct 14];23(1):43–7. Available from: https://www.tandfonline.com/doi/abs/10.1076/opge.23.1.43.2202

92. Ramírez-Castro JL, Pineda-Trujillo N, Valencia A V, Muñ Etó N CM, Botero O, Trujillo O, et al. Mutations in FOXL2 Underlying BPES (Types 1 and 2) in Colombian Families. Am J Med Genet [Internet]. 2002;113:47–51. Available from: https://onlinelibrary.wiley.com/doi/10.1002/ajmg.10741

93. Yamada T, Hayasaka S, Matsumoto M, Budu, Esa T, Hayasaka Y, et al. Heterozygous 17-bp deletion in the forkhead transcription factor gene, FOXL2, in a Japanese family with blepharophimosis-ptosis-epicanthus inversus syndrome. J Hum Genet [Internet]. 2001 Dec;46(12):733–6. Available from: https://www.nature.com/articles/jhg2001126

94. De Baere E, Van Roy N, Speleman F, Fukushima Y, De Paepe A, Messiaen L. Closing in on the BPES gene on 3q23: Mapping of a de Novo reciprocal translocation t(3;4)(q23;p15.2) Breakpoint within a 45-kb cosmid and mapping of three candidate genes, RBP1, RBP2, and β’-COP, distal to the breakpoint. Genomics. 1999 Apr 1;57(1):70–8.

95. Boccone L, Meloni A, Falchi AM, Usai V, Cao A. Blepharophimosis, ptosis, epicanthus inversus syndrome, a new case associated with de novo balanced autosomal translocation [46,XY,t(3;7)(q23;q32)]. Am J Med Genet. 1994;51(3):258–9.

96. Lawson CT, Toomes C, Fryer A, Carette MJM, Taylor GM, Fukushima Y, et al. Definition of the blepharophimosis, ptosis, epicanthus inversus syndrome critical region at chromosome 3q23 based on the analysis of chromosomal anomalies. Hum Mol Genet [Internet]. 1995 May 1 [cited 2022 Oct 14];4(5):963–7. Available from: https://academic.oup.com/hmg/article/4/5/963/708722

97. Wolstenholme J, Brown J, Masters KG, Wright C, English CJ. Blepharophimosis sequence and diaphragmatic hernia associated with interstitial deletion of chromosome 3 (46,XY,del(3)(q21q23)). J Med Genet [Internet]. 1994 [cited 2022 Oct 14];31(8):647. Available from: /pmc/articles/PMC1050030/?report=abstract

98. Costa T, Pashby R, Huggins M, Teshima IE. Deletion 3q in two patients with blepharophimosis-ptosis-epicanthus inversus syndrome (BPES). J Pediatr Ophthalmol Strabismus. 1998 Sep;35(5):271–6.

99. Jewett T, Rao PN, Weaver RG, Stewart W, Thomas IT, Pettenati MJ. Blepharophimosis, ptosis, and epicanthus inversus syndrome (BPES) associated with interstitial deletion of band 3q22: Review and gene assignment to the interface of band 3q22.3 and 3q23. Am J Med Genet. 1993;47(8):1147–50.

100. Ishikiriyama S, Goto M. Blepharophimosis sequence (BPES) and microcephaly in a girl with del(3) (q22.2q23): A putative gene responsible for microcephaly close to the BPES gene? Am J Med Genet. 1993;47(4):487–9.

101. Fujita H, Meng J, Kawamura M, Tozuka N, Ishii F, Tanaka N. Boy with a chromosome del(3)(q12q23) and blepharophimosis syndrome. Am J Med Genet. 1992;44(4):434–6.

102. Alvarado M, Bocian M, Walker AP. Interstitial deletion of the long arm of chromosome 3: Case report, review, and definition of a phenotype. Am J Med Genet. 1987;27(4):781–6.

103. Ye J, Shi X, He J jing, Zhang H na. Mutation analysis of FOXL2 gene in Chinese patients with blepharophimosis-ptosis-epicanthus inversus syndrome. Zhonghua Yan Ke Za Zhi [Internet]. 2011 Nov [cited 2022 Nov 7];47(11):1007–11. Available from: http://www.ncbi.nlm.nih.gov/pubmed/22336067

104. Wang Y, Wu Q, Cao W, Huang L, Liu W, Li C, et al. Clinical and genetic studies of 17 Han Chinese pedigrees and 31 sporadic patients with blepharophimosis-ptosis-epicanthus inversus syndrome. Mol Vis [Internet]. 2022 [cited 2022 Nov 24];28:352–8. Available from: http://www.ncbi.nlm.nih.gov/pubmed/36338666

105. Al V, Wj W, Bh S, Vincent AL, Watkins WJ, Sloan BH, et al. Blepharophimosis and bilateral Duane syndrome associated with a FOXL2 mutation. Clin Genet [Internet]. 2005 Dec 1 [cited 2022 Nov 8];68(6):520–3. Available from: https://onlinelibrary.wiley.com/doi/full/10.1111/j.1399-0004.2005.00527.x

106. Alao MJ, Lalèyè A, Lalya F, Hans C, Abramovicz M, Morice-Picard F, et al. Blepharophimosis, ptosis, epicanthus inversus syndrome with translocation and deletion at chromosome 3q23 in a black African female. Eur J Med Genet [Internet]. 2012 Nov [cited 2025 Aug 21];55(11):630–4. Available from: https://pubmed.ncbi.nlm.nih.gov/22906557/

107. Al-Awadi SA, Naguib KK, Farag TI, Teebi AS, Cuschieri A, Al-Othman SA, et al. Complex translocation involving chromosomes Y, 1, and 3 resulting in deletion of segment 3q23→q25. J Med Genet [Internet]. 1986 [cited 2025 Aug 21];23(1):91–2. Available from: https://pubmed.ncbi.nlm.nih.gov/3950944/

108. De Almeida JCC, Neto JBG, Jung M, Martins RR, Llerna JC. Another example favouring the location of BPES at 3q2. J Med Genet [Internet]. 1993 [cited 2025 Aug 21];30(1):86. Available from: https://pmc.ncbi.nlm.nih.gov/articles/PMC1016250/

109. Crisponi L, Uda M, Deiana M, Loi A, Nagaraja R, Chiappe F, et al. FOXL2 inactivation by a translocation 171 kb away: Analysis of 500 kb of chromosome 3 for candidate long-range regulatory sequences. Genomics [Internet]. 2004 May [cited 2025 Aug 21];83(5):757–64. Available from: https://pubmed.ncbi.nlm.nih.gov/15081106/

110. De Die-Smulders CEM, Engelen JJM, Donk JM, Fryns JP. Further evidence for the location of the BPES gene at 3q2. J Med Genet [Internet]. 1991 [cited 2025 Aug 21];28(10):725. Available from: https://pmc.ncbi.nlm.nih.gov/articles/PMC1017067/

111. Fukushima Y, Wakui K, Nishida T, Ueoka Y. Blepharophimosis sequence and de novo balanced autosomal translocation [46,XY,t(3;4)(q23;p15.2]: Possible assignment of the trait to 3q23. Am J Med Genet [Internet]. 1991 [cited 2025 Aug 21];40(4):485–7. Available from: https://pubmed.ncbi.nlm.nih.gov/1746616/

112. Praphanphoj V, Goodman BK, Thomas GH, Niel KM, Toomes C, Dixon MJ, et al. Molecular Cytogenetic Evaluation in a Patient with a Translocation (3;21) Associated with Blepharophimosis, Ptosis, Epicanthus Inversus Syndrome (BPES). Genomics [Internet]. 2000 Apr 1 [cited 2025 Aug 20];65(1):67–9. Available from: https://www.sciencedirect.com/science/article/pii/S0888754300961573?via%3Dihub

113. Schlade-Bartusiak K, Brown L, Lomax B, Bruyère H, Gillan T, Hamilton S, et al. BPES with atypical premature ovarian insufficiency, and evidence of mitotic recombination, in a woman with trisomy X and a translocation t(3;11)(q22.3;q14.1). Am J Med Genet A [Internet]. 2012 Sep [cited 2025 Aug 21];158 A(9):2322–7. Available from: https://pubmed.ncbi.nlm.nih.gov/22887799/

114. Warburg M, Bugge M, Brøndum-Nielsen K. Cytogenetic findings indicate heterogeneity in patients with blepharophimosis, epicanthus inversus, and developmental delay. J Med Genet [Internet]. 1995 [cited 2025 Aug 21];32(1):19–24. Available from: https://pubmed.ncbi.nlm.nih.gov/7897621/

115. Yang Y, Yang C, Zhu Y, Chen H, Zhao R, He X, et al. Intragenic and extragenic disruptions of FOXL2 mapped by whole genome low-coverage sequencing in two BPES families with chromosome reciprocal translocation. Genomics [Internet]. 2014 [cited 2025 Aug 21];104(3):170–6. Available from: https://pubmed.ncbi.nlm.nih.gov/25086333/

116. Yan YC, Zhou L, Fan JC. Identification and functional analyses of a novel FOXL2 pathogenic variant causing blepharophimosis, ptosis, and epicanthus inversus syndrome. Int J Ophthalmol [Internet]. 2023 [cited 2025 Oct 9];16(5):680–6. Available from: https://pubmed.ncbi.nlm.nih.gov/37206169/

117. Zhao M, Meng X, Wang J, Wang T. Novel FOXL2 variants in two Chinese families with blepharophimosis, ptosis, and epicanthus inversus syndrome. Front Genet [Internet]. 2024 Feb 12 [cited 2025 Oct 9];15:1343411. Available from: https://pmc.ncbi.nlm.nih.gov/articles/PMC10894958/

118. De Baere E, Beysen D, Oley C, Lorenz B, Cocquet J, De Sutter P, et al. FOXL2 and BPES: Mutational Hotspots, Phenotypic Variability, and Revision of the Genotype-Phenotype Correlation. Am J Hum Genet. 2003;72:478–87.

119. Shen Q, Zhao X, Ji Y, Chai P. Deletion of cis-regulatory Element in FOXL2 Promoter in a Chinese Family of Type II Blepharophimosis-ptosis-epicanthus Inversus Syndrome with Polydactyly. J Craniofac Surg [Internet]. 2023 Jan 1 [cited 2025 Sep 29];35(1):e52. Available from: https://pmc.ncbi.nlm.nih.gov/articles/PMC10749674/

120. Tzschach A, Kelbova C, Weidensee S, Peters H, Ropers HH, Ullmann R, et al. Blepharophimosis-ptosis-epicanthus inversus syndrome in a girl with chromosome translocation t(2;3)(q33;q23). Ophthalmic Genet [Internet]. 2008 Mar [cited 2025 Aug 21];29(1):37–40. Available from: https://pubmed.ncbi.nlm.nih.gov/18363172/

121. Tzschach A, Kelbova C, Weidensee S, Peters H, Ropers HH, Ullmann R, et al. Blepharophimosis-ptosis-epicanthus inversus syndrome in a girl with chromosome translocation t(2;3)(q33;q23). Ophthalmic Genet [Internet]. 2008 Mar [cited 2025 Aug 21];29(1):37–40. Available from: https://www.tandfonline.com/doi/abs/10.1080/13816810701867615

122. González-González C, García-Hoyos M, Calzón RH, Arroyo Díaz C, Fanego CG, Lorda Sánchez I, et al. Ophthalmic Genetics Microdeletion found by array-CGH in girl with Blepharophimosis syndrome and apparently balanced translocation t(3;15)(q23;q25). Ophthalmic Genet [Internet]. 2012 [cited 2025 Aug 21];33(2):107–10. Available from: https://www.tandfonline.com/action/journalInformation?journalCode=iopg20

123. Toomes C, Dixon MJ. Refinement of a translocation breakpoint associated with blepharophimosis-ptosis-epicanthus inversus syndrome to a 280-kb interval at chromosome 3q23. Genomics. 1998 Nov 1;53(3):308–14.

124. Dremsek P, Schachner A, Reischer T, Krampl-Bettelheim E, Bettelheim D, Vrabel S, et al. Retrospective study on the utility of optical genome mapping as a follow-up method in genetic diagnostics. J Med Genet [Internet]. 2025 [cited 2025 Aug 21];62:89–96. Available from: http://jmg.bmj.com/

125. Moumné L, Fellous M, Veitia RA. Deletions in the polyAlanine-containing transcription factor FOXL2 lead to intranuclear aggregation. Hum Mol Genet [Internet]. 2005 Dec 1 [cited 2022 Oct 11];14(23):3557–64. Available from: https://academic.oup.com/hmg/article/14/23/3557/559445

126. Dipietromaria A, Benayoun BA, Todeschini AL, Rivals I, Bazin C, Veitia RA. Towards a functional classification of pathogenic FOXL2 mutations using transactivation reporter systems. Hum Mol Genet [Internet]. 2009 Sep 1 [cited 2022 Oct 12];18(17):3324–33. Available from: https://academic.oup.com/hmg/article/18/17/3324/2527401

127. Beysen D, Moumné L, Veitia R, Peters H, Leroy BP, De Paepe A, et al. Missense mutations in the forkhead domain of FOXL2 lead to subcellular mislocalization, protein aggregation and impaired transactivation. Hum Mol Genet [Internet]. 2008 Jul [cited 2022 Oct 11];17(13):2030–8. Available from: https://pubmed.ncbi.nlm.nih.gov/18372316/

128. Caburet S, Demarez A, Moumné L, Fellous M, De Baere E. A recurrent polyalanine expansion in the transcription factor FOXL2 induces extensive nuclear and cytoplasmic protein aggregation. J Med Genet [Internet]. 2004 [cited 2022 Oct 11];41:932–6. Available from: www.jmedgenet.com

129. Laissue P, Lakhal B, Benayoun BA, Dipietromaria A, Braham R, Elghezal H, et al. Functional evidence implicating FOXL2 in non-syndromic premature ovarian failure and in the regulation of the transcription factor OSR2. J Med Genet [Internet]. 2009 [cited 2022 Nov 24];46:455–7. Available from: http://jmg.bmj.com/

130. Moumné L, Dipietromaria A, Batista F, Kocer A, Fellous M, Pailhoux E, et al. Differential aggregation and functional impairment induced by polyalanine expansions in FOXL2, a transcription factor involved in cranio-facial and ovarian development. Hum Mol Genet [Internet]. 2008 Apr 1 [cited 2022 Oct 11];17(7):1010–9. Available from: https://academic.oup.com/hmg/article/17/7/1010/869933

131. Pailhoux E, Vigier B, Chaffaux S, Servel N, Taourit S, Furet JP, et al. A 11.7-kb deletion triggers intersexuality and polledness in goats. Nature Genetics 2001 29:4 [Internet]. 2001 Nov 19 [cited 2022 Oct 26];29(4):453–8. Available from: https://www.nature.com/articles/ng769z

132. Simon R, Lischer HEL, Pieńkowska-Schelling A, Keller I, Häfliger IM, Letko A, et al. New genomic features of the polled intersex syndrome variant in goats unraveled by long-read whole-genome sequencing. Anim Genet [Internet]. 2020 Jun 1 [cited 2023 Mar 31];51(3):439–48. Available from: https://onlinelibrary.wiley.com/doi/full/10.1111/age.12918

133. Vaiman D, Pailhoux E, Schibler L, Oustry A, Chaffaux S, Cotinot C, et al. Genetic mapping of the polled/intersex locus (PIS) in goats. Theriogenology [Internet]. 1997 Jan 1 [cited 2025 Sep 29];47(1):103–9. Available from: https://www.sciencedirect.com/science/article/pii/S0093691X96003445

134. Shi F, Ding S, Zhao S, Han M, Zhuang Y, Xu T, et al. A piggyBac insertion disrupts Foxl2 expression that mimics BPES syndrome in mice. Hum Mol Genet [Internet]. 2014 Jul 15 [cited 2022 Oct 26];23(14):3792–800. Available from: https://academic.oup.com/hmg/article/23/14/3792/558425

135. Bashamboo A, Eozenou C, Jorgensen A, Bignon-Topalovic J, Siffroi JP, Hyon C, et al. Loss of Function of the Nuclear Receptor NR2F2, Encoding COUP-TF2, Causes Testis Development and Cardiac Defects in 46,XX Children. Am J Hum Genet [Internet]. 2018 Mar 1 [cited 2025 Oct 9];102(3):487–93. Available from: https://pubmed.ncbi.nlm.nih.gov/29478779/

136. Méjécase C, Nigam C, Moosajee M, Bladen JC. The Genetic and Clinical Features of FOXL2-Related Blepharophimosis, Ptosis and Epicanthus Inversus Syndrome. Genes (Basel) [Internet]. 2021 Mar 1 [cited 2025 Aug 28];12(3):364. Available from: https://pmc.ncbi.nlm.nih.gov/articles/PMC7998575/

