## Supplemental Text 1 for "Variant curation of the largest compendium of *FOXL2* coding and non-coding sequence and structural variants in BPES"

**Supplementary text**

All variants were classified using our in-house ACMG/AMP-based framework, which evaluates 7 evidence categories: (1) population data, (2) genotype/phenotype, (3) public databases, (4) computational/predictive data, (5) functional data, (6) segregation data, and (7) allelic data.

For **population data**, gnomAD v4.1.0 was queried. Variants absent from gnomAD supported PM2. PS4 was considered when the same rare variant was identified in multiple unrelated patients with BPES (supporting if in 1, moderate if in ≥2).

For **genotype/phenotype**, *FOXL2* is the only known disease gene for BPES, which has typical characteristics. PP4 was therefore always applicable. The strength of evidence was adjusted: PP4 moderate when detailed clinical description or photographs were available, and PP4 strong when additional biochemical confirmation (e.g., FSH and/or AMH levels indicating POI) was provided.

For **public databases**, PP5 and BP6 were not applied in accordance with Biesecker *et al.* (2018), except when variants were submitted to ClinVar without published evidence, in which case the ClinVar classification was considered.

For **computational and predictive data**, REVEL <0.4 supported BP4 and >0.7 supported PP3; intermediate values were not scored. PM5 was used for missense changes affecting codons with a known pathogenic variant. PM4 was applied to in-frame insertions and deletions, particularly polyalanine expansions. The criterion was considered supporting when a single amino acid was inserted or deleted, and moderate when more than one amino acid was affected. Null variants were evaluated under PVS1, considering that *FOXL2* is a single-exon gene.

For **functional data**, published studies were considered (Caburet *et al.,* 2004; Moumné *et al.*, 2008; Dipietromaria *et al.*, 2009; Li *et al.*, 2021). Pathogenic effects demonstrated at mRNA or protein level supported PS3. Missense variants in the forkhead domain were assigned PM1.

For **segregation data**, PP1 was applied when segregation with BPES was demonstrated. PM6 was applied when variants were likely *de novo* without confirmed parentage. For **allelic data**, no applicable criteria were identified in this cohort.

All criteria were combined according to ACMG/AMP guidelines to reach a final variant classification.
